# Beyond Gambling-Related Cognitive Distortions: Irrational Thinking and the Role of Compulsivity in Problem Gambling

**DOI:** 10.64898/2025.12.22.25342819

**Authors:** Jennifer Molitor, Jan Peters

## Abstract

**Background and aims:** Gambling-related cognitive distortions (GRCD) are closely linked to problem gambling symptom severity (PGSI) and are associated with superstitious and delusional ideation, as well as indirectly with conspiracy beliefs. Although conceptually distinct, these different belief domains share core features (e.g., erroneous beliefs about causal structure) and may be related to compulsivity. The latent factor structure underlying these belief domains is poorly understood. This preregistered study examined a potentially shared latent structure underlying GRCD, superstitious and delusional ideation, and conspiracy beliefs, and links with PGSI and a transdiagnostic symptom dimension previously linked to addictive and compulsive psychopathology.

**Methods:** Participants with previous gambling experience (N = 491) were recruited via Prolific © and completed measures assessing each belief domain, along with dimensions of compulsive behavior and intrusive thought, anxious-depression, and social withdrawal. Several factor analytic models were compared to determine the optimal latent structure.

**Results:** PGSI was significantly correlated with all irrational belief domains. Model comparison favored a bifactor model comprising a general factor accounting for over half of the variance (55%, closely aligned with superstitious content) and domain-specific factors related to GRCD and conspiracy beliefs. The association between compulsivity and PGSI was partially mediated by GRCDs, suggesting compulsivity may affect PGSI both directly and indirectly via a modulation of GRCDs.

**Conclusions:** Findings confirm that problem gambling is associated with irrational beliefs beyond GRCDs and support a dimensional neurocognitive model in which general and domain-specific belief components are linked to compulsivity. This suggests shared mechanisms underlying the formation and persistence of irrational beliefs across domains, with unique features specific to GRCD.

## INTRODUCTION

Decision-making under uncertainty relies on cognitive heuristics to infer predictability from incomplete or noisy information (Gigerenzer & Gaissmaier, 2011; Tversky & Kahneman, 1974). While generally adaptive, these mechanisms can produce systematic errors (*cognitive distortions*), especially in inherently stochastic domains like gambling (Hahn & Warren, 2009; Matute et al., 2015). Gambling disorder (GD) is characterized by persistent gambling despite adverse consequences (American Psychiatric Association, 2013). With an estimated prevalence of around 1.4 % (Tran et al., 2024), GD represents a significant public health concern. Maladaptive gambling-related cognitive distortions (GRCDs) are strongly linked to GD severity ((Brooks & Clark, 2022; Buen & Flack, 2022; Goodie & Fortune, 2013; Thurm et al., 2023) and are major treatment targets (Fortune & Goodie, 2012).

GRCDs may emerge partly from the exposure to gambling (Peters, 2025; Redish et al., 2007), and include perceived control, unrealistic expectations of gratification, and interpretative biases, such as attributing wins to skill rather than chance (Goodie & Fortune, 2013; Raylu & Oei, 2004; Toneatto, 1999). Prominent examples are the gamblers’ fallacy and the illusion of control (Goodie & Fortune, 2013; Jacobsen et al., 2007; Langer, 1975), both reflecting attempts to impose causal structure on stochastic events. The gamblers’ fallacy represents a predictive control illusion, an overestimation of one’s ability to predict random outcomes (Raylu & Oei, 2004) such as expecting a result to be more or less likely based on prior events (Cowan, 1969; Tversky & Kahneman, 1971). The illusion of control is the belief that chance-based outcomes can be influenced through personal action (Langer, 1975), such as wearing a "lucky" shirt, pressing slot machine buttons in a specific sequence, or praying for success. Both distortions are not exclusive to gambling but likely reflect broader cognitive biases (Hahn & Warren, 2009; Stefan & David, 2013).

GRCDs are robustly associated with conceptually related belief domains (e.g., Brooks & Clark, 2022) that attribute causality to non-empirical forces, such as superstitious, magical, or delusional ideation. Recent work links the illusion of control more strongly to superstitious beliefs than to agency perception (O. Griffiths et al., 2019; Klusowski et al., 2021; Monson et al., 2024). Superstition includes beliefs in luck, destiny, or astrology, and it is strongly associated with GRCD and GD severity (Brooks & Clark, 2022; Fluke et al., 2014; M. D. Griffiths & Bingham, 2005; Joukhador et al., 2004; Leonard & Williams, 2019). Delusional ideation also refers to erroneous and persistent beliefs (American Psychiatric Association, 2022), although often self-referential, including ideas like being targeted or chosen by a higher power. Subclinical levels are common and correlate with both GRCD and GD severity (Abdollahnejad et al., 2014, 2015; Brooks & Clark, 2022). Conspiracy beliefs extend erroneous attributions to malevolent groups (Frenken & Imhoff, 2022). No study to date has directly linked conspiracy beliefs to GRCDs or GD, but individuals endorsing conspiracy beliefs are more likely to hold other irrational beliefs, including paranormal beliefs, delusional ideation, and demonstrate higher psychopathology (Brotherton & French, 2014; Dagnall et al., 2015; dos Reis et al., 2024; Moulding et al., 2016). Moreover, both conspiracy and superstitious beliefs are linked to heightened pattern perception in noise, such as chaotic artwork, global events, and coin tosses (van Prooijen et al., 2018), a tendency that is particularly pronounced under low control (Whitson & Galinsky, 2008).

Taken together, these domains share the tendency to infer erroneous causal structure and persist despite contradictory evidence. It remains unclear whether observed associations reflect a shared latent structure or domain-specific processes. We address this gap by exploring the latent structure across belief domains, providing a basis for assessing whether more general cognitive traits contribute to such distorted beliefs.

### Compulsive Behavior and Intrusive Thought

One trait that might contribute to irrational beliefs is compulsivity. GD involves compulsivity symptoms that are also central to substance use disorders and obsessive-compulsive disorder (OCD; Everitt & Robbins, 2005; Gillan & Robbins, 2014), such as rigid, repetitive thoughts and behaviors that persist despite negative consequences (Gillan et al., 2016; Robbins et al., 2012).

Compulsive behavior and intrusive thought (CIT; (Boldt et al., 2024; Friedemann et al., 2024; Gillan et al., 2016; Wise & Dolan, 2020) is a transdiagnostic construct that is linked to deficits in goal-directed control, thereby potentially linking OCD, SUDs, and behavioral addictions, such as gambling. Figee et al. (2016) described several neurobiological mechanisms related to compulsivity, including: reduced dopaminergic signaling in the ventral striatum and increased reliance on negative reinforcement via limbic circuits; diminished serotonergic modulation in prefrontal regions, contributing to cognitive and behavioral rigidity; and a shift from goal-directed to habitual responding, reflecting dysregulated ventral-to-dorsal frontostriatal dynamics.

CIT and underlying neurocognitive mechanisms may contribute to cognitive distortions across belief domains. For example, compulsive symptoms, particularly in OCD, are linked to magical, superstitious, and delusional ideation (Bortolon & Raffard, 2015; Einstein & Menzies, 2004a, 2004b, 2006; Frost et al., 1993; Mauzay et al., 2016; Mauzay & Cuttler, 2018; Sica et al., 2002). Moreover, conspiracy beliefs are associated with reduced cognitive flexibility (Sambol et al., 2024). In this view, GRCDs may not only reflect gambling-specific biases, but might arise from broader cognitive vulnerabilities, particularly those linked to CIT, e.g. due to excessive reliance on prior beliefs and an insensitivity to feedback. This dovetails with findings of behavioral inflexibility and diminished goal-directed control in problem gambling (Brands, Knauth, et al., 2025; Bruder et al., 2021; van Timmeren et al., 2018; Wyckmans et al., 2019). Generally, the relationship between cognitive flexibility, goal-directed control, and compulsivity is well established (Fineberg et al., 2015; Gillan et al., 2016). However, the only study directly investigating links between the transdiagnostic CIT dimension and gambling severity observed a non-significant positive association in a small sample (N = 38; Friedemann et al., 2024), underscoring the need for larger-scale investigations.

Therefore, the present preregistered study (https://doi.org/10.17605/OSF.IO/YX5BD) examined whether CIT, relative to the transdiagnostic dimensions of anxious-depression (AD) and social-withdrawal (SW), is reliably associated with problem gambling symptoms in a large sample of individuals participating in gambling. We hypothesized CIT to be associated with problem gambling symptom severity. We further investigated the latent structure across belief domains (as outlined above) and how the resulting latent belief factors relate to both PGSI and CIT.

## METHOD

### Participants

A total of 506 participants with lifetime gambling experience (≥ 1 occasion) were recruited via the online platform Prolific ©. After applying data quality criteria (e.g., attention checks, response latency), 15 participants (2.96%) were excluded, yielding a final sample of N = 491 (27.1% female, 72.1% male, 0.8% diverse, *M_age_* = 32.69, *SD_age_* = 9.91, *Range_age_* = 18–74). Sample characteristics are summarized in Table 1.

**Table 1.** Sample Characteristics and Demographics.

| Variable | n (%) | M (SD) / Mdn |
| --- | --- | --- |
| Final sample size | 491 | — |
| Female | 133 (27.1%) | — |
| Male | 354 (72.1%) | — |
| Diverse | 4 (0.8%) | — |
| Age (years) | — | $M = 32.69$ ($SD = 9.91$) |
| Migration background | 214 (43.6%) | — |
| Subjective SES | — | $Mdn = 6$ |
| Monthly income (€) | — | $Mdn = 2,200$ |
| Higher education entrance qualification | 394 (80.2%) | — |
| Gambling activities | | $M = 3.43$ ($SD = 2.34$) |
| Gambling frequency (past 12 months) |  |  |
| No gambling | 49 (10%) |  |
| < 1 x per month | 225 (45.8%) |  |
| ≈ 1 x per month | 63 (12.8%) |  |
| 2-3 x per month | 87 (17.7%) |  |
| ≥ 1 x per week | 67 (13.6%) |  |

*Note.* SES = Socioeconomic status on a 0–10 scale, with higher values indicating higher subjective SES. Gambling activities = average number of different gambling activities per participant.

*A priori* power analysis (G*Power; Faul et al., 2009) was based on the smallest relevant correlation previously reported (*r* = .17; Brooks & Clark, 2022), with a two-tailed *α* of .05 and a power of .95. This resulted in a required sample size of N = 439. To account for potential exclusions, we increased our preregistered target sample size by 15% to N = 505.

### Procedure

The survey was administered online via LimeSurvey (Limesurvey GmbH, n.d.), with all measures presented in randomized order. Completion took approximately 20 minutes, and participants received on average €4.15 compensation.

### Measures

Scales lacking a validated German reference were translated using an iterative forward-backward translation procedure.

#### Demographics

Participants reported age, gender, educational level, income, socioeconomic status (Hoebel et al., 2015), and migration background.

#### Gambling Related Cognitive Distortions

The 23-item *Gambling-Related Cognitions Scale* (GRCS; Raylu & Oei, 2004) assesses five subdomains: inability to stop gambling, gambling expectations, illusion of control, predictive control, and interpretive bias. Responses were given on a 7-point scale. Cronbach’s *α* = .91.

#### Superstition

The 25-item *Superstitious Belief Questionnaire* (SBQ; Griffiths et al., 2019) captures paranormal, supernatural, magical, and superstitious beliefs on a 0–4 scale. The original preamble, referencing “evidence for psi phenomena”, was replaced (see Supplement). Cronbach’s *α* = .95.

#### Delusional Ideation

The 21-item *Peters et al. Delusions Inventory* (PDI; Lincoln, 2024) measures proneness to delusional thinking. For each endorsed experience, participants rated strength, distress, and preoccupation on a 5-point Likert scale. Cronbach’s *α* = .72.

#### Conspiracy Thinking

The 15-item *Generic Conspiracist Belief Scale* (GCB; Brotherton et al., 2013) measures endorsement of common conspiracy themes on a 5-point scale. Cronbach’s *α* = .94.

#### Gambling Behavior

Engagement in various gambling activities (e.g., slot machines, sports betting) during the past 12 months was rated on a 5-point scale (“did not participate at all”, “less than once a month”, “once a month”, “2-3 times a month”, “at least weekly”).

#### Gambling Symptoms

Gambling Symptoms in the past 12 months were assessed using the 9-item *Problem Gambling Severity Index* (PGSI; Ferris & Wynne, 2001), rated on a 4-point scale. The PGSI yields a classification into non-problem, low-risk, moderate-risk, and problem gambling. Cronbach’s *α* = .91.

#### Transdiagnostic Dimensions

CIT, anxious-depression (AD), and social-withdrawal (SW) were assessed using a set of previously validated items from existing scales (Boldt et al., 2024; Gillan et al., 2016; Wise & Dolan, 2020). Composite scores were computed using published factor weights (Wise & Dolan, 2020). Cronbach’s *α* CIT = .87; AD = .94; SW = .90.

### Statistical Analysis

Preregistered analyses included an exploratory factor analysis (EFA) on GRCS, PDI, SBQ, and GCB items to identify latent belief factors. Maximum likelihood estimation (MLE) with oblimin rotation was applied. Factor retention followed preregistered criteria, including scree plot based on Cattell’s criterion (Cattell, 1966) and Cattell–Nelson–Gorsuch test (CNG test; Gorsuch & Nelson, 1981). Items were assigned to the factor with the highest loading, applying a threshold of ≥ .30. In cases of relevant cross-loadings (<.20 difference), assignments were guided by theoretical considerations.

Based on results, we conducted additional non-preregistered confirmatory factor analyses (CFA) to test competing orthogonal, hierarchical, and bifactor models, and evaluated model fit based on: Root Mean Square Error of Approximation (RMSEA), Comparative Fit Index (CFI), Trucker-Lewis Index (TLI), Bayesian Information Criterion (BIC), and Standardized Root Mean Square Residual (SRMR).

In addition, we conducted pre-registered bivariate correlation analyses between the extracted belief factors and transdiagnostic symptom dimensions (CIT, AD, SW), as well as with PGSI. Finally, we tested a non-preregistered mediation model using structural equation modeling (SEM) with bootstrapped confidence intervals to determine the degree to which the gambling-specific beliefs factor mediated the association between CIT and PGSI.

All analyses were conducted in R (version 4.4.0) using the *psych* (2.5.3; Revelle, 2025) and *lavaan* (0.6.19; Rosseel, 2012) packages. Item distributions were examined for normality, and Box-Cox transformations were applied. Items were z-standardized prior to analysis. Detailed diagnostics and transformation parameters are provided in the Supplementary Methods.

### Ethics

The study was conducted in accordance with the Declaration of Helsinki and approved by the ethics committee of the University of Cologne. All subjects provided informed consent.

## RESULTS

### Association between PGSI and irrational belief domains

PGSI scores (M = 2.34, SD = 4.05) yielded a classification of participants as follows: 45.2% non-problematic gambling, 27.1% low-risk gambling, 20.8% moderate-risk gambling, and 6.9% probable problem gambling.

As expected, GRCS scores increased with PGSI severity (*r* = .56, *p* < .001, 95% CI [.50, .63]), from M = 37.83 (SD = 13.43; non-problem) to M = 75.32 (SD = 23.41; probable problem gambling) (see Figure 1a). Scores on PDI (delusional ideation, Figure 1b), SBQ (superstition, Figure 1c), and GCB (conspiracy beliefs, Figure 1d) were all positively associated with PGSI scores, and all intercorrelations were significant (see Figure 1e).

**Figure 1.**
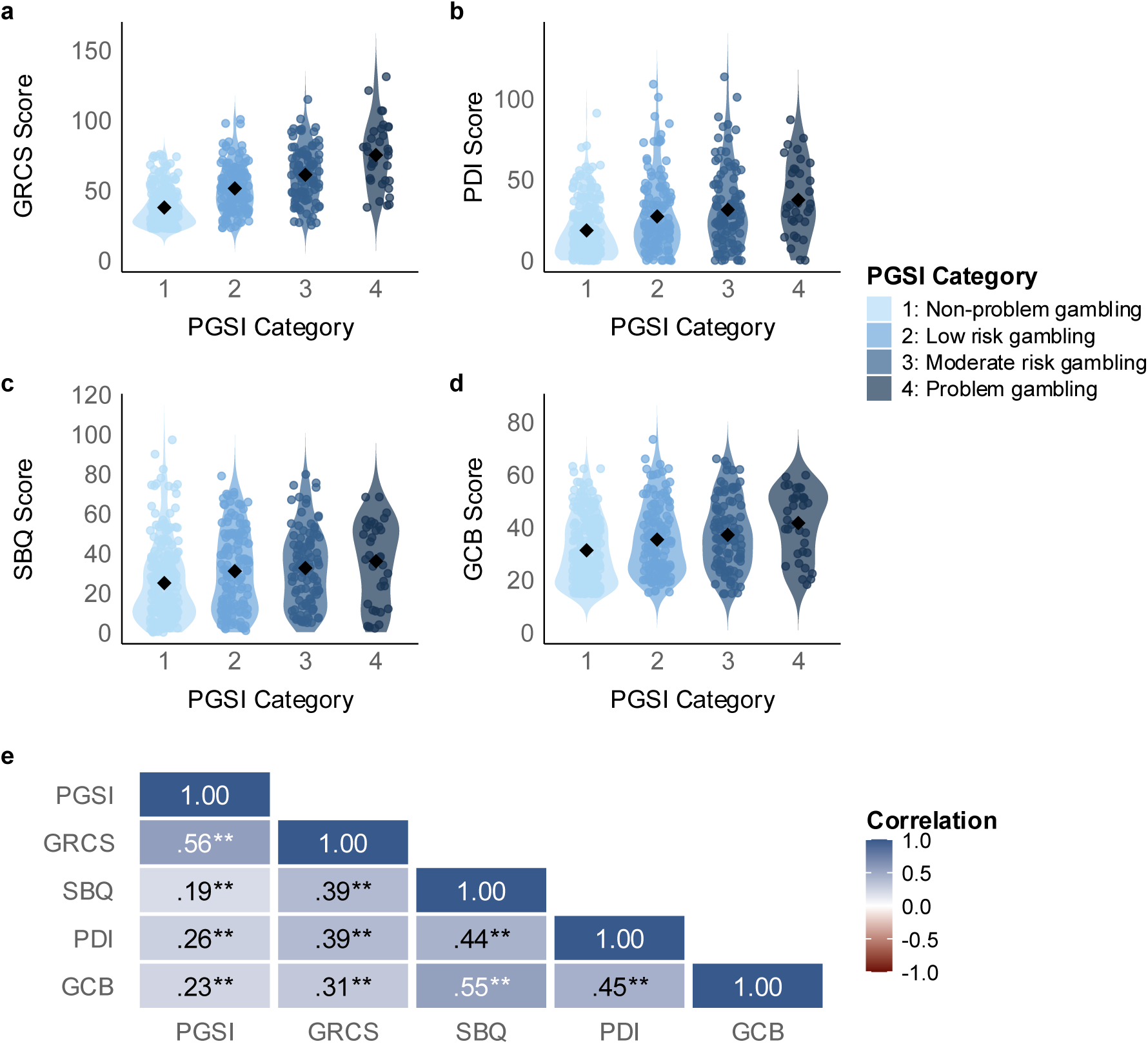
(a-d) Distribution of (a) GRCS, (b) PDI, (c) SBQ, and (d) GCB scores by PGSI category. Dots represent individual participants; black diamonds denote group means. (e) Pearson correlations among PGSI, GRCS, SBQ, PDI, GCB (all p < .01). Shading indicates correlation strength. GRCS = Gambling Related Cognitions Scale (Raylu & Oei, 2004); PDI = Peters Delusion Inventory (Lincoln, 2024); SBQ = Superstitious Belief Questionnaire (O. Griffiths et al., 2019); GCB = Generic Conspiracist Beliefs Scale (Brotherton et al., 2013); PGSI = Problem Gambling Symptom Severity Index (Ferris & Wynne, 2001).

### EFA

An EFA on GRCS (M = 48.98, SD = 20.12), PDI (M = 24.90, SD = 21.78), SBQ (M = 28.82, SD = 20.23), and GCB (M = 34.21, SD = 13.21) items showed high sampling adequacy (MSA = .94), and significant Bartlett’s test (*p* < .05). The scree plot suggested a four-factor solution, whereas the CNG test indicated three.

The three-factor model (see Figure 2) explained 34.1% of variance (RMSEA = .044, TLI = .818, and BIC = –13,442). Factor 1 was defined predominantly by SBQ items, with minor contributions from three PDI and one GRCS items, reflecting a superstition dimension. Factor 2 consisted primarily of GRCS items, representing GRCDs. Factor 3 contained all GCB items, corresponding to conspiracy beliefs (CB). Cross-loadings were minimal (<.30), except for a single GRCS item (.35) loading on both superstition and GRCD. Weak loadings of PDI items (<.30) were diffusely distributed across factors.

**Figure 2.**
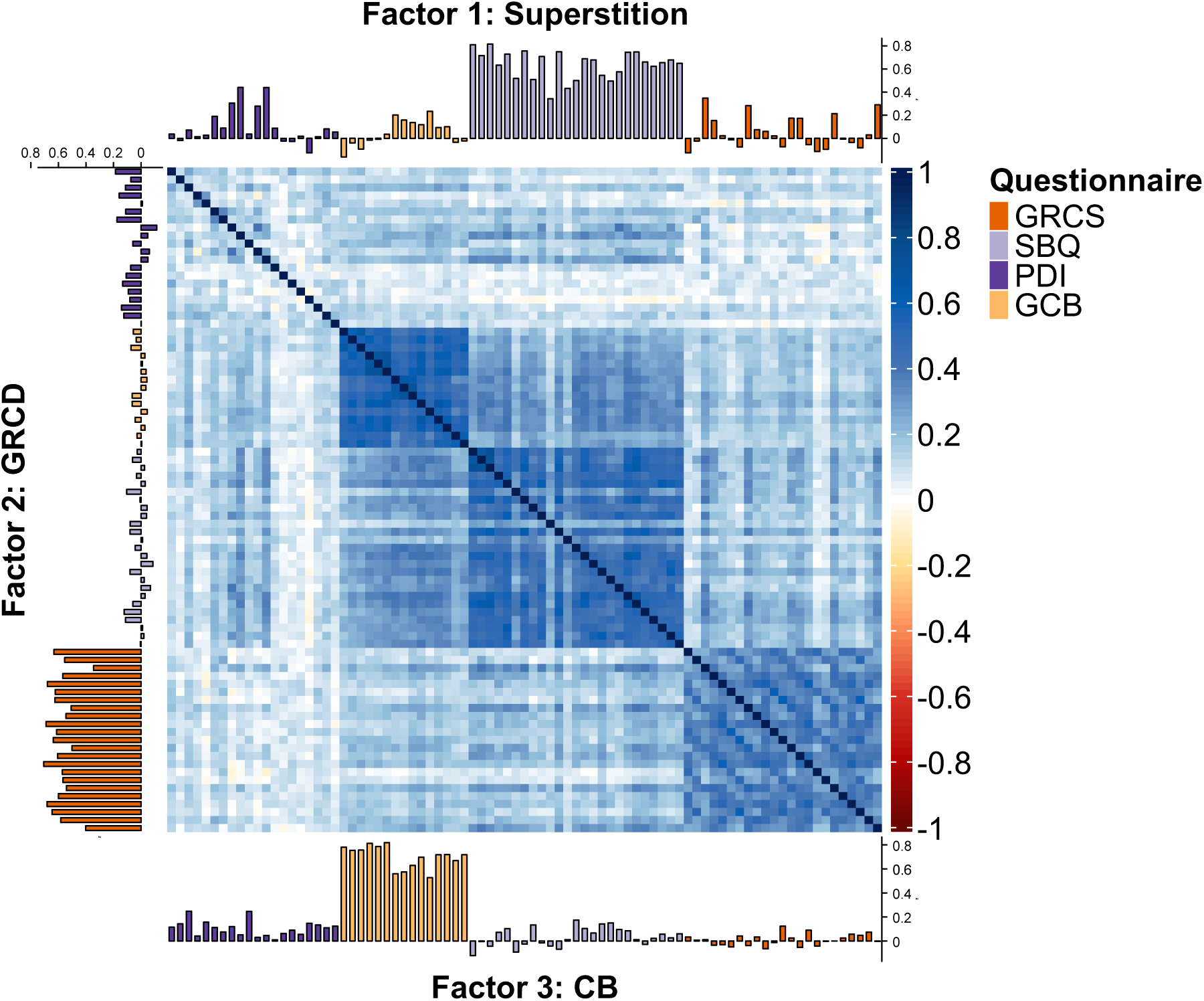
Visual representation of the three-factor EFA solution, showing item-level correlations and factor loadings. GRCD = Gambling-related cognitive distortions; CB = Conspiracy beliefs; GRCS = Gambling Related Cognitions Scale (Raylu & Oei, 2004); PDI = Peters Delusion Inventory (Lincoln, 2024); SBQ = Superstitious Belief Questionnaire (O. Griffiths et al., 2019); GCB = Generic Conspiracist Beliefs Scale (Brotherton et al., 2013).

The four-factor model accounted for 35.7% of variance (RMSEA = .039, TLI = .851, BIC = –13,635), aligning more closely with original scales: Factor 1 (SBQ), Factor 2 (GRCS), Factor 3 (GCB), and Factor 4 (PDI). However, the fourth factor (PDI) was weakly defined with loadings < .30 and diffuse cross-loadings, explaining only 1.9% of the variance. Given the minimal added value and weaker definition, the three-factor model (Figure 2) was retained.

### CFA Models

Three non-preregistered CFA models were tested (Figure 3): (a) an orthogonal (independence of domains), (b) a hierarchical (three domain-specific factors loading onto a higher-order factor), and (c) a bifactor model (general and domain-specific factors).

**Figure 3.**
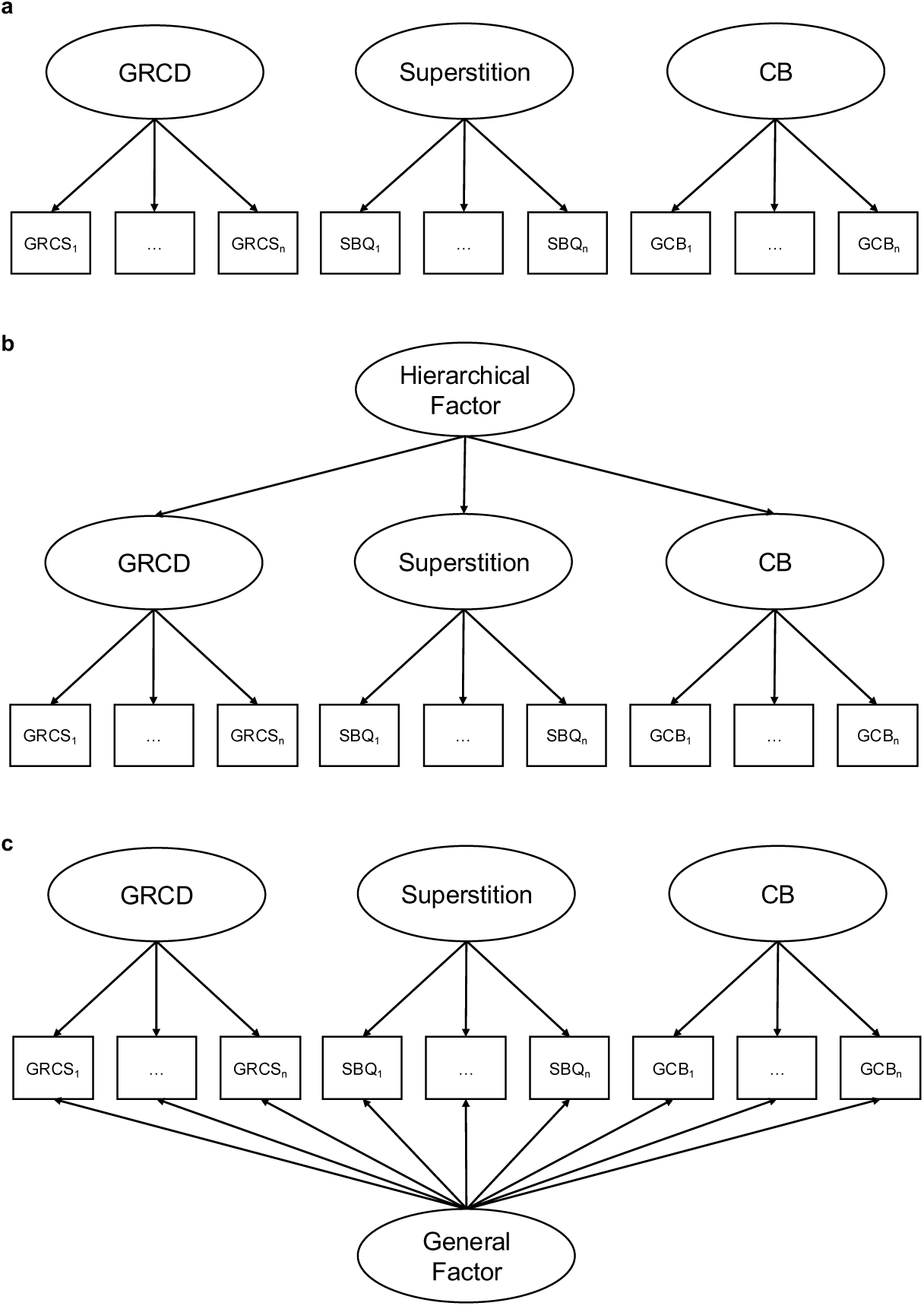
Schematic representation of (a) an orthogonal three-factor model (b) a hierarchical model, and (c) a bifactor model. GRCD = Gambling-related cognitive distortions; CB = Conspiracy beliefs; GRCS = Gambling Related Cognitions Scale; SBQ = Superstitious Belief Questionnaire; GCB = Generic Conspiracist Beliefs Scale

#### a. Orthogonal Model

The three-factor CFA with orthogonal latent variables corresponds to the assumption that GRCD, superstitious, and conspiracy beliefs represent independent domains. Model fit was the poorest among all tested models (see Table 2), indicating that the independence assumption was unacceptable.

**Table 2.** CFA model comparison of fit indices.

| **Model** | **$\chi^2$ (df)** | **RMSEA**<br>**[90% CI]** | **CFI** | **TLI** | **SRMR** | **AIC** | **BIC** |
| --- | --- | --- | --- | --- | --- | --- | --- |
| Orthogonal | 5218.97<br>(2015) | .057<br>[.055, .059] | .820 | .814 | .179 | 76,149 | 76,695 |
| Hierarchical | 4941.07<br>(2012) | .054<br>[.053, .056] | .835 | .829 | .064 | 75,878 | 76,436 |
| Bifactor | 4328.78<br>(1914) | .051<br>[.049, .053] | .863 | .856 | .093 | 74,085 | 74,782 |

#### b. Hierarchical Model

A hierarchical CFA was estimated with three first-order factors (GRCD, Superstition, and CB) loading onto a higher-order factor. All domain factors loaded significantly, with standardized estimates of .86 (Superstition), .68 (CB), and .51 (GRCD). Model fit was improved relative to the orthogonal model (see Table 2), indicating that a general tendency toward distorted beliefs accounted for shared variance, while meaningful domain-specific variance remained.

#### c. Bifactor Model

To separate general from domain-specific variance, a bifactor structure was estimated. As bifactor models cannot be fully specified a priori, an EFA using maximum likelihood extraction and bifactor rotation was conducted. The general factor explained 21% of the variance, and three domain-specific factors emerged: gambling (8%), conspiracy (6%), and superstition (4%). The bifactor model demonstrated the best fit and interpretability among all tested models (Table 2). Omega coefficients confirmed excellent total reliability (*ω*_*t*_ = .97), with 55% of reliable variance attributable to the general factor. Gambling (*ω* = .82) and conspiracy (*ω* = .64) factors showed unique reliability, whereas the unique variance for a superstition-specific factor was negligible (*ω* = .04). SBQ items loaded primarily on the general factor (except item SBQ10). Domain-specific loadings for superstition items were marginal. GRCS items loaded mainly on the gambling-specific factor, although several items showed loadings on the general factor. All GCB items loaded on both the general and the conspiracy-specific factor. From the PDI, only items PDI9 and PDI12 (telepathy and witchcraft) exceeded the threshold on the general factor; none loaded sufficiently on any domain-specific factor.

Item-specific variance decomposition (Figure 4) showed that most GRCD items were domain specific. However, illusion of control and predictive control items also shared variance with the general factor, with one item (“Praying helps me win”) fully explained by this dimension. Superstition items were almost entirely absorbed by the general factor, while conspiracy items showed mixed contributions, with domain-specific variance accounting for the larger share.

**Figure 4.**
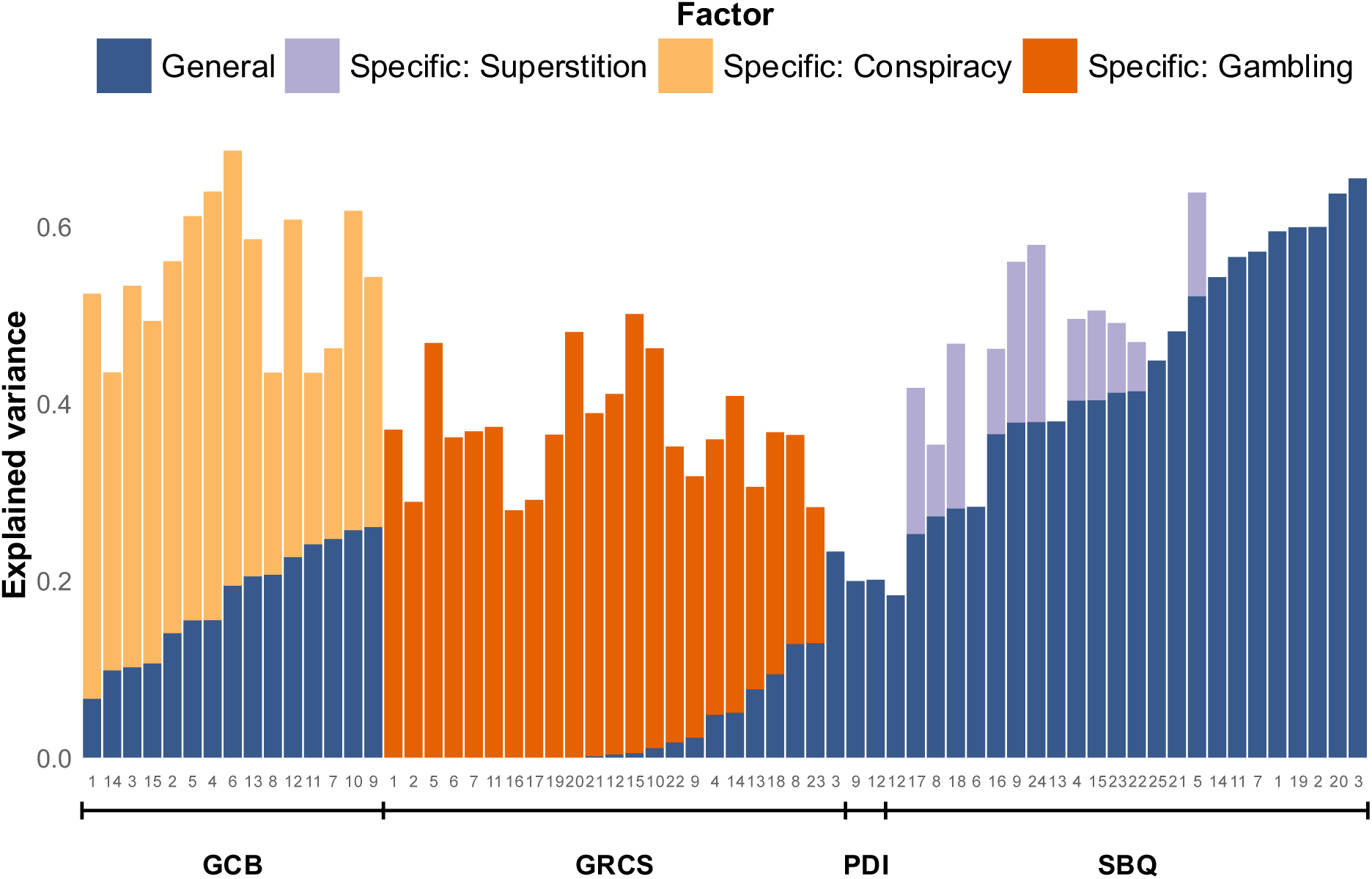
Explained variance for each item in the bifactor model, partitioned into general and domain-specific factors (gambling, conspiracy, superstition). Most variance in GRCD items is domain-specific; conspiracy showed mixed variance proportions; variance of superstition items was mainly absorbed by the general factor. GRCS = Gambling Related Cognitions Scale (Raylu & Oei, 2004); PDI = Peters Delusion Inventory (Lincoln, 2024); SBQ = Superstitious Belief Questionnaire (O. Griffiths et al., 2019); GCB = Generic Conspiracist Beliefs Scale (Brotherton et al., 2013).

### Correlation Analysis

We next examined Spearman correlations between PGSI, transdiagnostic dimensions (CIT, AD, SW), and general and gambling-specific latent factors from the best-fitting bifactor model. In line with our preregistered hypothesis, PGSI scores were positively correlated with CIT (medium effect size, Figure 5a). AD and SW showed small, significant positive correlations with PGSI (Figure 5f).

**Figure 5.**
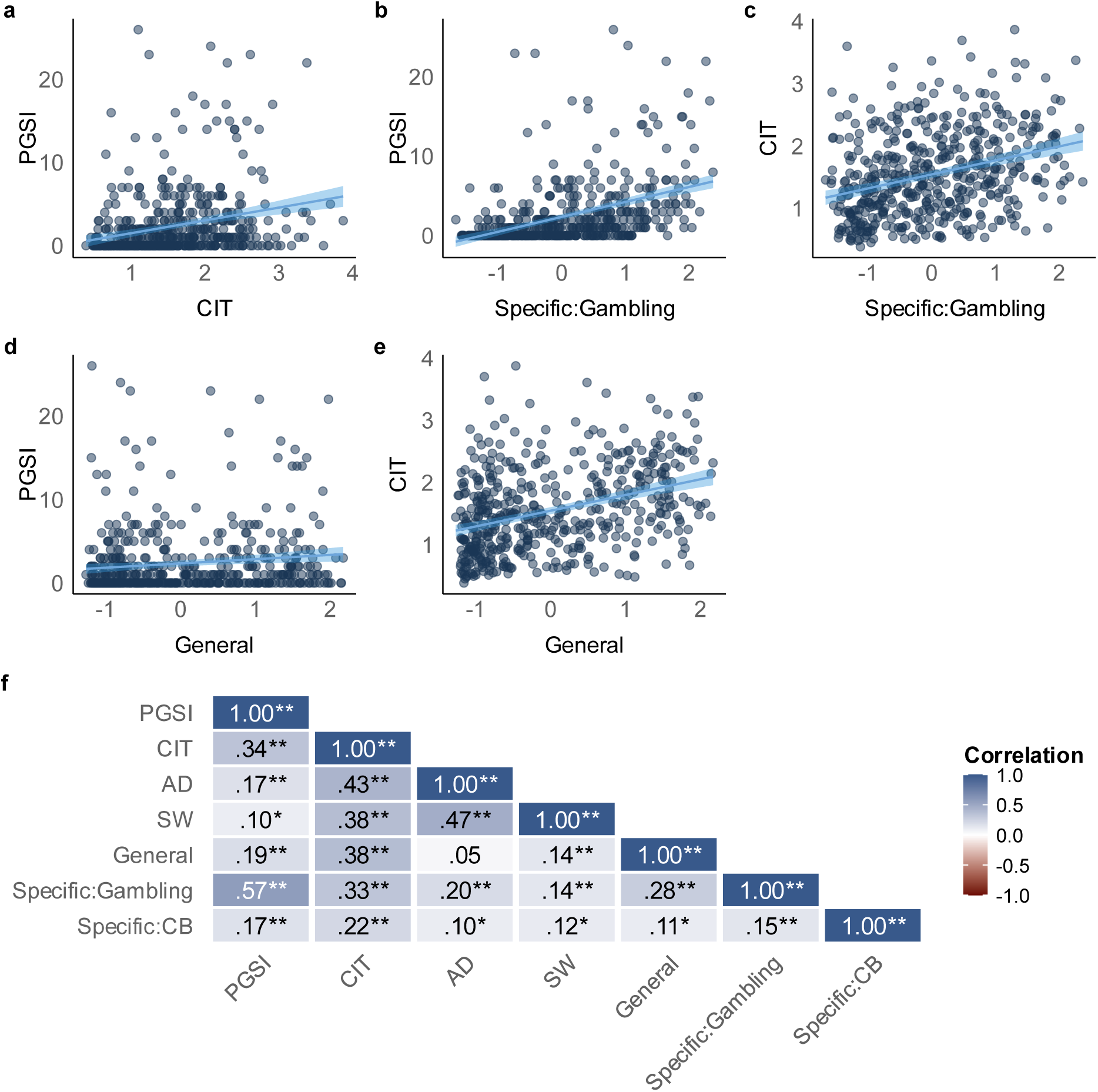
(a-e) Scatterplots, illustrating the associations between PGSI, CIT, General and Specific: Gambling. Dark-blue dots represent individual participants; blue regression line represents the linear fit; shaded blue band represents the 95% confidence interval. (f) Pearson correlations among PGSI, CIT, AD, SW, General, Specific: Gambling, and Specific: CB. Significance is indicated as p < .05* and p < .01**. Shading indicates correlation strength. PGSI = Problem Gambling Symptom Severity Index (Ferris & Wynne, 2001); CIT = Compulsive Behavior and Intrusive Thought, AD = Anxious-Depression, SW = Social-Withdrawal (Gillan et al., 2016; Wise & Dolan, 2020); General = General Factor; Specific: Gambling = Gambling-specific Factor; Specific: CB = Conspiracy-specific Factor.

CIT was positively correlated with the gambling-specific and general factors (Figure 5c, e, medium effect sizes). The gambling-specific factor showed a medium effect size correlation with PGSI (Figure 5b), whereas the correlation with the general factor was small (Figure 5d).

### Mediation Analysis

Based on this pattern of associations, we next carried out a non-preregistered mediation analysis (Figure 6) to test the idea that CIT may constitute a vulnerability factor that predisposes individuals to develop GRCDs during exposure to gambling, which may in turn be linked to PGSI. All individual paths were statistically significant: CIT predicted gambling-specific distortions, gambling-specific distortions predicted PGSI, and CIT showed a significant direct effect on PGSI.

**Figure 6.**
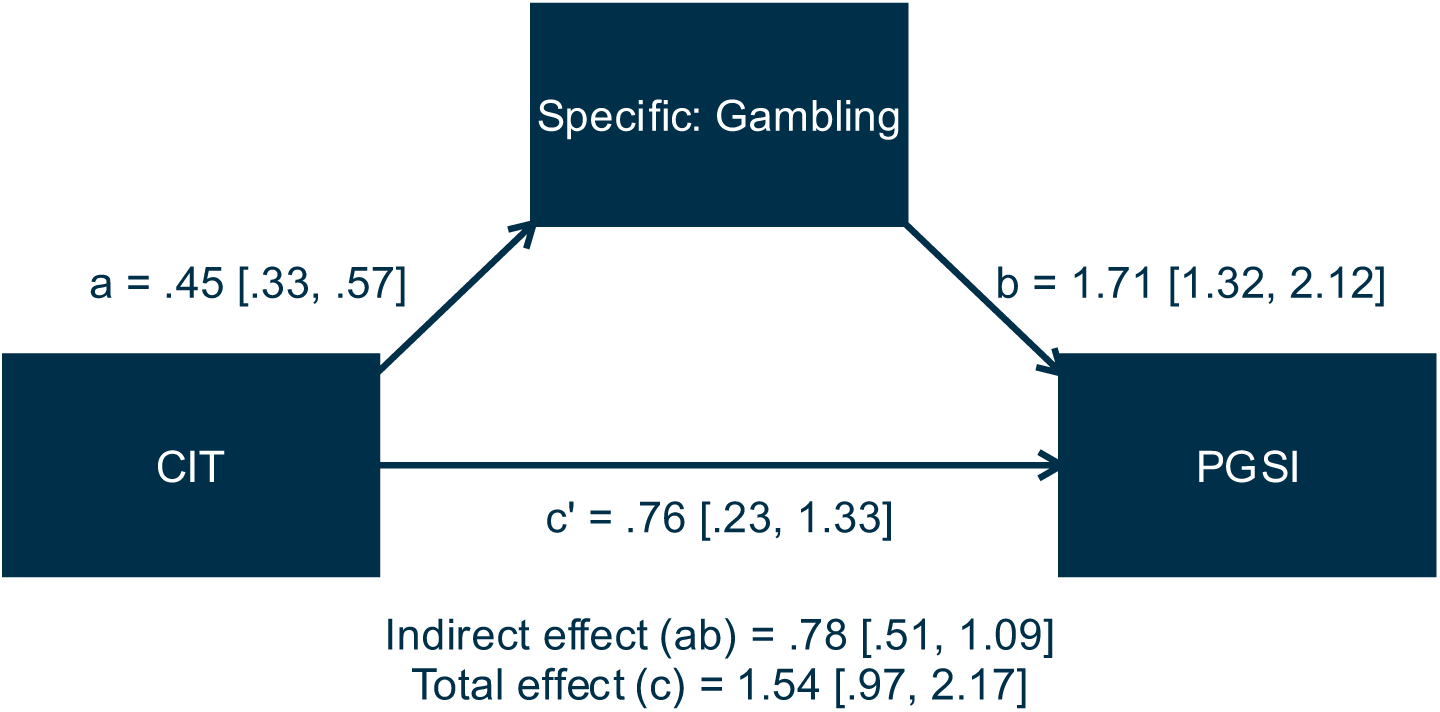
Mediation model, illustrating the indirect effect of CIT on PGSI through the gambling-specific cognitive distortions factor (Specific: Gambling), with unstandardized regression coefficients and 95% percentile bootstrap confidence intervals (5,000 samples). CIT = Compulsive behavior and intrusive thought (Gillan et al., 2016; Wise & Dolan, 2020); PGSI = Problem gambling severity index (Ferris & Wynne, 2001).

The indirect effect was significant (SE = .15, *z* = 5.18, *p* < .001), as was the total effect (SE = .30, *z* = 5.22, *p* < .001). Gambling-specific cognitions mediated 50.5% of the total effect (SE = .12, *z* = 4.38, *p* < .001), indicating partial mediation.

## DISCUSSION

This study examined associations between problem gambling symptom severity (PGSI) and a range of irrational belief domains in a large online sample of individuals engaging in gambling. PGSI was positively associated with gambling-related cognitive distortions (GRCDs), superstitious, delusional, and conspiratorial beliefs, suggesting that cognitive distortions in gambling may reflect a broader pattern of irrational thinking, rather than a set of isolated gambling-specific beliefs. We also confirmed that previously identified transdiagnostic dimensions related to compulsive behavior and intrusive thought (CIT), anxious-depressive thoughts (AD), and social-withdrawal (SW; Boldt et al., 2024; Friedemann et al., 2024; Gillan et al., 2016; Wise & Dolan, 2020) exhibit small (SW) to moderate (CIT) positive correlations with PGSI.

Factor analyses supported a bifactor structure comprising a general factor related to irrational beliefs, and three domain-specific factors: gambling, conspiracy beliefs (CB), and superstition. The general factor captured most variance in superstition and partial variance in gambling and conspiracy items. The unique variance attributable to a separate superstition-specific factor was negligible. Within the gambling domain, illusion of control and predictive control items loaded partially on the general factor, consistent with prior interpretations that the illusion of control reflects superstition, rather than beliefs grounded in perceived control (M. D. Griffiths & Bingham, 2005; Klusowski et al., 2021; Monson et al., 2024). Other GRCS subscales were exclusively accounted for by domain-specific variance, highlighting their contextual dependence. CB items showed both general and domain-specific variance.

We also examined associations between PGSI and CIT. Replicating the non-significant finding of Friedemann et al. (2024) in a much larger sample, our data provide first robust evidence linking PGSI and CIT, complementing earlier findings associating CIT with compulsivity and addiction-related phenotypes (Gillan et al., 2016; Seow & Gillan, 2020). Importantly, a mediation analysis revealed that the gambling-specific factor partially mediated this relationship, accounting for approximately half of the total effect, while the direct effect remained significant. CIT may thus contribute to problem gambling via two routes: indirectly through a modulation of gambling-specific beliefs, and directly via potential behavioral mechanisms. This is consistent with the role of compulsivity in promoting persistent behavior despite adverse consequences (Everitt & Robbins, 2005; Gillan et al., 2016). These results are also consistent with the idea that CIT may predispose individuals to develop GRCDs during exposure to gambling (Peters, 2025; Redish et al., 2007), although longitudinal studies are required to test this causal pathway more directly.

Both the general and the gambling-specific factors exhibited positive correlations with CIT, suggesting that compulsive tendencies are associated with irrational thinking across contexts. The general factor was only weakly associated with PGSI, whereas the gambling-specific factor showed a strong correlation. Additionally, AD demonstrated weak associations with both PGSI and the gambling-specific factor, consistent with the well-documented role of comorbid depressive symptoms in GD (Karlsson & Håkansson, 2018; Lorains et al., 2011; Quigley et al., 2015).

The general factor might reflect culturally shared or commonly experienced cognitive distortions such as the gambler’s fallacy (Farmer et al., 2017; Hahn & Warren, 2009), as well as other distortions shaped through social learning and/or exposure to cultural narratives. In contrast, gambling-specific distortions may require exposure to specific experiences to emerge, as outlined in recent theoretical models (Peters, 2025; Redish et al., 2007). Specific gambling design features could foster the emergence of such beliefs (Peters, 2025; Yücel et al., 2018). In line with our mediation results, exposure to gambling may provide a contextual framework through which general tendencies to endorse cognitive distortions (such as CIT) are expressed. However, this hypothesis requires further direct empirical validation. Superstition may represent a specific manifestation of associative learning processes. It has been linked to the cognitive system s attempt to detect (hidden) associations and infer cause-and-effect bonds (Daprati et al., 2019), reflecting an increased drive to generate explanations for perceived patterns (Beck & Forstmeier, 2007). This perspective emphasizes that superstition is not merely the result of faulty learning, but also of an active cognitive bias toward hypothesis formation, combined with an absence (or disregard) of feedback. (Abbott & Sherratt, 2011) demonstrated that superstitious behavior is more likely when the perceived benefits outweigh potential costs and when prior beliefs support the superstition, with both uncertainty and individual learning history modulating its likelihood. These findings suggest that superstition may arise from an adaptive tendency to detect causal structure under uncertainty, which can, however, lead to erroneous inferences when feedback is ambiguous or sparse.

At a mechanistic level, compulsive symptomatology, characterized by reduced goal-directed control, cognitive inflexibility, and reduced feedback sensitivity, may facilitate distortions across domains. Individuals with higher levels of compulsive neurocognition may be particularly prone to maintaining irrational beliefs through habitual cognitive and behavioral patterns, even in the face of disconfirming evidence. Consistent with this, impairments in model-based decision-making, i.e. a shift towards habitual, less flexible decision strategies, have been robustly associated with problem gambling (Brands, Knauth, et al., 2025; Bruder et al., 2021; van Timmeren et al., 2018; Wyckmans et al., 2019). Persistent cognitive distortions could reflect habitual patterns of thought emerging from associative learning processes that underlie stimulus-response habits (Cushman & Morris, 2015; Gillan et al., 2016).

Alternatively, compulsive behaviors themselves may elicit post-hoc rationalizations, leading to distorted cognitions that serve to justify or make sense of one’s actions (Gillan et al., 2014; Gillan & Robbins, 2014). Both mechanisms may contribute to the observed associations, though our design precludes causal interpretations. Together, these perspectives highlight how general compulsive tendencies and domain-specific learning experiences could jointly contribute to gambling-related cognitive distortions.

### Limitations and Future Directions

The interpretation of our findings is subject to several limitations. The PDI differs structurally from the other instruments due to its gating procedure, which may have reduced variance and attenuated loadings. However, cross-loadings suggest that delusional ideation was not empirically separable from other belief domains in this dataset. Furthermore, the factor analyses were exploratory, and the overall model fit was moderate. The structure should be regarded as preliminary and requires validation in confirmatory studies with independent samples.

Although the general factor correlated with CIT based on self-report, future research could incorporate behavioral measures of goal-directed control (e.g., model-based learning tasks; (Brands, Knauth, et al., 2025; Brands, Mathar, et al., 2025; Bruder et al., 2021; Gillan et al., 2016; Voon et al., 2017) to test the links to compulsivity and behavioral flexibility more directly. Generally, the present study is limited by its cross-sectional nature. Longitudinal data would be required to assess if the gambling-specific distortion factor predicts outcomes such as escalation from recreational to problem gambling or treatment response, and to directly test the idea of CIT as a predisposing vulnerability factor for the development of GRCDs following gambling exposure.

Further limitations concern the sample and data acquisition mode. First, we collected data from a large online sample of individuals with gambling experience. As a result, the proportion of participants exhibiting moderate-risk gambling and probable problem gambling were 20.8% and 6.9%, respectively, and conclusions are thus limited to individuals not clinically diagnosed with GD. Likewise, the proportion of female participants substantially decreased with increasing PGSI levels (reflecting well-known prevalence differences (Tran et al., 2024)), but this precluded us from systematically investigating potential gender effects. Second, we used established criteria to ensure data quality, and all instruments exhibited good to excellent internal consistency in this data set. Nonetheless, potential data quality concerns related to online studies more generally need to be considered. Third, we focused solely on self-report. Future studies would benefit from using task-based measures (Voon et al., 2017) to obtain insights into specific cognitive and computational mechanisms.

Finally, we have explicitly focused on irrational belief domains in problem gambling. However, similar mechanisms might contribute to other forms of addictive behaviors(Billieux et al., 2020; Bodi et al., 2021; Brooks & Clark, 2019; Davis, 2001; Flayelle et al., 2023; King & Delfabbro, 2016; Marino & Spada, 2017)

## Conclusion

The present study demonstrates that problem gambling symptom severity is associated with multiple domains of irrational thinking, including superstitious, delusional, and conspiratorial thinking. Factor analysis revealed a general factor, capturing culturally shared distortions primarily linked to superstition, and gambling- and conspiracy-specific factors, reflecting domain-specific cognitive distortions. Both the general and the gambling-specific factors were significantly associated with a previously established transdiagnostic compulsivity factor (CIT), and the gambling-specific factor partially mediated the link between CIT and problem gambling symptom severity. Compulsive neurocognition may provide a common foundation for the persistence of irrational beliefs and may determine experience-dependent belief expression following gambling exposure. Our results highlight the importance of integrating models of irrational belief formation with frameworks of compulsivity neurocognition. Future research may help to delineate how cultural exposure, domain-specific experience, and compulsive neurocognitive traits jointly shape the emergence and persistence of irrational beliefs in gambling and related behaviors.

## Data Availability

All data produced in the present study are available upon request to the authors.

