## Supplementary Information for "Beyond Gambling-Related Cognitive Distortions: Irrational Thinking and the Role of Compulsivity in Problem Gambling"

### Supplementary Method

#### Preamble SBQ

To avoid misleading participants about the existence of “recent evidence for psi-phenomena the preamble was replaced with: People repeatedly experience phenomena that cannot be explained by known physical or biological theories. As a first step, we want to find out what people think about some of these topics. You will be presented with a series of statements and asked to rate how strongly you agree or disagree with each one. It is very important that you answer as honestly as possible.

#### Transdiagnostic Questionnaire

**Table 1**

*Transdiagnostic Questionnaire Items*

| Scale | German Version | Nr. | Content |
|---|---|---|---|
| **Self-rating Depression Scale (SDS; Zung, 1965),** | Collegium Internationale Psychiatriae Scalarum, CIPS (Ed.), 1986 | 11 | My mind is as clear as it used to be. (R) |
| | | 12 | I find it easy to do the things I used to. (R) |
| | | 13 | I am restless and can’t keep still/sleep. |
| | | 14 | I feel hopeful about the future. (R) |
| | | 16 | I find it easy to make decisions. (R) |
| | | 17 | I feel that I am useful and needed. (R) |
| | | 18 | My life is pretty full. (R) |
| | | 20 | I still enjoy the things I used to do. (R) |

| | | | |
|---|---|---|---|
| **Obsessive-Compulsive Inventory (OCI; Foa et al., 2002)** | Gönner et al., 2007 | 1 | I have saved up so many things that they get in the way. |
| | | 2 | I check things more often than necessary. |
| | | 4 | I feel compelled to count while I am doing things. |
| | | 6 | I find it difficult to control my own thoughts. |
| | | 7 | I collect things I don't need. |
| | | 9 | I get upset if others change the way I have arranged things. |
| | | 11 | I sometimes have to wash or clean myself simply because I feel contaminated. |
| | | 12 | I am upset by unpleasant thoughts that come into my mind against my will. |
| | | 13 | I avoid throwing things away because I am afraid I might need them later. |
| | | 16 | I feel that there are good and bad numbers. |
| | | 18 | I frequently get nasty thoughts and have difficulty in getting rid of them. |
| | | 2 | I get things done during the day. (R) |

| | | | |
|---|---|---|---|
| **Apathy Evaluation Scale (AES; Marin et al., 1991)** | Lueken et al., 2006 | 7 | I approach life with intensity. (R) |
| | | 17 | I have initiative. (R) |
| | | 18 | I have motivation. (R) |
| **Eating Attitude Test (EAT; Garner et al., 1982)** | Meermann & Vandereycken, 2019 | 1 | I am terrified about being overweight. |
| | | 11 | I am preoccupied with a desire to be thinner. |
| | | 12 | I think about burning up calories when I exercise. |
| | | 14 | I am preoccupied with the thought of having fat on my body. |
| **Baratt Impulsiveness Scale (BIS-11; Patton et al., 1995)** | | 1 | I plan tasks carefully. (R) |
| | | 6 | I have “racing” thoughts |
| | | 9 | I concentrate easily. (R) |
| | | 13 | I plan for job security. (R) |
| | | 14 | I say things without thinking. |
| | | 15 | I like to think about complex problems. (R) |
| | | 17 | I act “on impulse”. |
| | | 20 | I am a steady thinker. (R) |
| | | 22 | I buy things on impulse. |
| | | 25 | I spend or charge more than I earn. |
| | | 26 | I often have extraneous thoughts when thinking. |

| | | | |
|---|---|---|---|
| **State Trait Anxiety Inventory (STAI; Spielberger et al., 1983)** | | 1 | I feel pleasant. (R) |
| | | 3 | I feel satisfied with myself. (R) |
| | | 5 | I feel like a failure. |
| | | 8 | I feel that difficulties are piling up so that I cannot overcome them. |
| | | 9 | I worry too much over something that really doesn't matter. |
| | | 10 | I am happy. (R) |
| | | 12 | I lack self-confidence. |
| | | 13 | I feel secure. (R) |
| | | 16 | I am content. (R) |
| | | 19 | I am a steady person (R) |
| | | 20 | I get in a state of tension or turmoil as I think over my recent concerns and interests. |
| **Liebowitz Social Anxiety Scale (LSAS; Liebowitz, 1987)** | Mitte et al., 2007 | 2 | Participating in small groups. |
| | | 7 | Going to a party. |
| | | 8 | Working while being observed. |
| | | 10 | Calling someone you don't know very well. |
| | | 11 | Talking with people you don't know very well. |
| | | 12 | Meeting strangers. |
| | | 14 | Entering a room when others are already seated. |

| | | | |
|---|---|---|---|
| | | 15 | Being the center of attention. |
| | | 16 | Speaking up at a meeting. |
| | | 18 | Expressing a disagreement or disapproval to people you don't know very well. |
| | | 20 | Giving a report to a group. |
| | | 23 | Giving a party. |
| | | 24 | Resisting a high pressure salesperson. |
| **Alcohol Use Disorder Identification Test (AUDIT; Saunders et al., 1993)** | Dybek et al., 2006 | 1 | How often do you have a drink containing alcohol. |

*Note.* R = Reverse

**Supplementary Results**

Normality of GRCS, PDI, SBQ and GCB items was assessed at both the multivariate and univariate levels. The Henze-Zirkler test indicated significant multivariate non-normality (HZ = 1.008, $p < .001$). Additionally, Anderson-Darling tests revealed significant deviations from univariate normality for all items ($p < .001$; Table 2), with some skewness values exceeding 5 and markedly elevated kurtosis, particularly for PDI items. Due to violations of normality assumptions, a Box-Cox transformation was applied (Table 2). This transformation selects an optimal lambda value (λ) to minimize skewness and improve normality. Since this method requires strictly positive values, all variables containing zeros were shifted by adding one while preserving the original data structure. To ensure comparability across scales, all items were subsequently z-standardized.

To assess sampling adequacy, the Kaiser-Meyer-Olkin (KMO) measure was computed. One item (PDI21) was excluded due to a low MSA value of 0.59. The final analysis included 83 items, with an overall MSA of 0.94. Bartlett's test was significant ($p$ < .05), indicating that the correlation structure was suitable for factor analysis.

Given that Pearson correlations are sensitive to non-normality, Spearman's rank correlation was used to compute the correlation matrix. This approach is more robust under distributional violations and thus better suited for the present data.

**Table 2**

Item diagnostics with Box-Cox parameters

| Item | Lambda | Shift | Mean | SD | Skew | Kurtosis |
|---|---|---|---|---|---|---|
| PDI1 | 0.28893360 | 1 | 3.9694501 | 3.6350173 | 0.2716336 | 1.800727 |
| PDI2 | -9.63903899 | 1 | 0.3279022 | 1.4751048 | 4.9344594 | 28.708833 |
| PDI3 | -0.27763471 | 1 | 3.5906314 | 4.1641612 | 0.5437600 | 1.733337 |
| PDI4 | -6.35792645 | 1 | 0.5702648 | 2.1344243 | 3.8168837 | 16.751031 |
| PDI5 | -10.48779775 | 1 | 0.3380855 | 1.6449538 | 5.2057648 | 30.629319 |
| PDI6 | -2.87075050 | 1 | 1.1160896 | 2.6179845 | 2.2047240 | 6.569395 |
| PDI7 | -1.26954805 | 1 | 1.9409369 | 3.1445733 | 1.3214539 | 3.413572 |
| PDI8 | -5.15017610 | 1 | 0.6537678 | 2.1001169 | 3.1894736 | 12.255548 |
| PDI9 | -3.75589590 | 1 | 0.8146640 | 2.1750601 | 2.6148843 | 8.652471 |
| PDI10 | -2.28513326 | 1 | 1.5539715 | 3.3610372 | 1.9338396 | 5.275498 |
| PDI11 | -10.19474940 | 1 | 0.3279022 | 1.5092962 | 4.6808087 | 24.136537 |

| Item | Lambda | Shift | Mean | SD | Skew | Kurtosis |
|---|---|---|---|---|---|---|
| PDI12 | -5.29429421 | 1 | 0.6415479 | 2.1135677 | 3.2971573 | 12.826121 |
| PDI13 | -3.25271370 | 1 | 1.1181263 | 2.9092213 | 2.4858809 | 7.920641 |
| PDI14 | -3.42235976 | 1 | 1.0061100 | 2.6729872 | 2.6184639 | 8.762298 |
| PDI15 | -1.17308941 | 1 | 2.2505092 | 3.6050382 | 1.2598413 | 3.128726 |
| PDI16 | -3.97578582 | 1 | 0.8391039 | 2.3751645 | 2.8463899 | 10.154177 |
| PDI17 | -2.26064459 | 1 | 1.6659878 | 3.5757751 | 1.8297549 | 4.700175 |
| PDI18 | -3.60410815 | 1 | 0.9979633 | 2.7161640 | 2.5718508 | 8.165784 |
| PDI19 | -5.48469886 | 1 | 0.6048880 | 2.0787301 | 3.7007923 | 16.539392 |
| PDI20 | -11.44683343 | 1 | 0.2871690 | 1.4189024 | 5.3374138 | 32.694176 |
| PDI21 | -12.73830587 | 1 | 0.2830957 | 1.5357307 | 5.8198665 | 37.743862 |
| GCB1 | 0.15575752 | 1 | 2.5254582 | 1.2860569 | 0.2950639 | 1.839196 |
| GCB2 | -0.40409211 | 1 | 2.2219959 | 1.1924956 | 0.5994145 | 2.220660 |
| GCB3 | 0.48270441 | 1 | 2.6313646 | 1.2423455 | 0.1363392 | 1.835381 |
| GCB4 | -0.32803081 | 1 | 2.3279022 | 1.2553622 | 0.5907227 | 2.176121 |
| GCB5 | -0.49524438 | 1 | 2.2627291 | 1.2802327 | 0.5813318 | 2.032984 |
| GCB6 | -0.46666450 | 1 | 2.2016293 | 1.1926978 | 0.6181084 | 2.219361 |
| GCB7 | -3.63440252 | 1 | 1.5519348 | 0.8983269 | 1.6279598 | 4.940333 |
| GCB8 | -1.00055401 | 1 | 2.0448065 | 1.1858126 | 0.8837669 | 2.706025 |
| GCB9 | -2.17224549 | 1 | 1.7250509 | 0.9896215 | 1.2163016 | 3.546043 |

| Item | Lambda | Shift | Mean | SD | Skew | Kurtosis |
|---|---|---|---|---|---|---|
| GCB10 | -0.74560539 | 1 | 2.0651731 | 1.1265936 | 0.7461704 | 2.457669 |
| GCB11 | -1.86350684 | 1 | 1.8268839 | 1.0975717 | 1.1435227 | 3.216977 |
| GCB12 | -0.82009526 | 1 | 2.0896130 | 1.1746098 | 0.8469697 | 2.666939 |
| GCB13 | -0.24641141 | 1 | 2.3625255 | 1.2900486 | 0.4964390 | 1.993509 |
| GCB14 | 0.98789044 | 1 | 2.9857434 | 1.2656371 | -0.1304215 | 1.919678 |
| GCB15 | 1.66682432 | 1 | 3.3910387 | 1.2428740 | -0.5100186 | 2.200598 |
| SBQ1 | -1.45083984 | 1 | 0.8024440 | 1.1621910 | 1.2012475 | 3.128157 |
| SBQ2 | -3.00156291 | 1 | 0.4806517 | 0.8766427 | 1.8512056 | 5.706627 |
| SBQ3 | -1.89253679 | 1 | 0.6782077 | 1.0739559 | 1.4479620 | 3.888731 |
| SBQ4 | 0.48628549 | 1 | 1.6211813 | 1.3120631 | 0.1205966 | 1.711026 |
| SBQ5 | -0.26426669 | 1 | 1.2606925 | 1.3421209 | 0.5923781 | 1.960646 |
| SBQ6 | -3.54913981 | 1 | 0.4073320 | 0.8062683 | 2.3135524 | 8.314948 |
| SBQ7 | -1.17345264 | 1 | 0.8431772 | 1.1525700 | 1.1569923 | 3.122583 |
| SBQ8 | 0.43989629 | 1 | 1.6130346 | 1.3205415 | 0.1671450 | 1.741001 |
| SBQ9 | -0.11580484 | 1 | 1.3014257 | 1.3181535 | 0.4715486 | 1.808852 |
| SBQ10 | 0.11660591 | 1 | 1.5519348 | 1.4648436 | 0.3168386 | 1.643956 |
| SBQ11 | -1.28102685 | 1 | 0.7902240 | 1.1004573 | 1.2125170 | 3.336980 |
| SBQ12 | 0.30894217 | 1 | 1.4521385 | 1.2613770 | 0.2393334 | 1.749704 |
| SBQ13 | 0.14487836 | 1 | 1.5071283 | 1.3870576 | 0.3526917 | 1.752039 |

| Item | Lambda | Shift | Mean | SD | Skew | Kurtosis |
|---|---|---|---|---|---|---|
| SBQ14 | -1.48650000 | 1 | 0.7739308 | 1.1265862 | 1.2493685 | 3.363865 |
| SBQ15 | 0.17260389 | 1 | 1.4501018 | 1.3259786 | 0.3074842 | 1.710593 |
| SBQ16 | 0.04274153 | 1 | 1.2525458 | 1.1803530 | 0.4706460 | 1.993290 |
| SBQ17 | 1.01993660 | 1 | 2.1608961 | 1.4187442 | -0.2983944 | 1.755469 |
| SBQ18 | -0.35905033 | 1 | 1.3116090 | 1.4521791 | 0.6485239 | 1.967486 |
| SBQ19 | -1.14638485 | 1 | 0.7841141 | 1.0469743 | 1.1335661 | 3.163310 |
| SBQ20 | -1.58666054 | 1 | 0.7189409 | 1.0778037 | 1.4078699 | 3.923696 |
| SBQ21 | -1.71762840 | 1 | 0.6578411 | 1.0066284 | 1.5288061 | 4.507541 |
| SBQ22 | -0.57396886 | 1 | 0.9592668 | 1.1131728 | 0.9511524 | 2.890797 |
| SBQ23 | -0.08635126 | 1 | 1.2729124 | 1.2740981 | 0.4630195 | 1.833038 |
| SBQ24 | 0.58028624 | 1 | 1.7006110 | 1.3038325 | 0.1016985 | 1.837846 |
| SBQ25 | 0.25609595 | 1 | 1.4663951 | 1.3079003 | 0.2596846 | 1.731244 |
| GRCS1 | 0.39168308 | 1 | 2.9551935 | 1.6181460 | 0.1622508 | 1.803036 |
| GRCS2 | -10.12783808 | 1 | 1.2830957 | 0.8051489 | 3.6951226 | 18.608298 |
| GRCS3 | -5.59008590 | 1 | 1.4989817 | 1.1237233 | 2.4471321 | 8.286905 |
| GRCS4 | -2.36525654 | 1 | 1.8085540 | 1.2020425 | 1.3950851 | 3.927771 |
| GRCS5 | -0.50774728 | 1 | 2.6700611 | 1.7868853 | 0.5865939 | 1.921698 |
| GRCS6 | -0.96979712 | 1 | 2.3665988 | 1.6200354 | 0.8915056 | 2.521551 |
| GRCS7 | -3.87767511 | 1 | 1.6008147 | 1.1497645 | 2.3050168 | 8.285552 |

| Item | Lambda | Shift | Mean | SD | Skew | Kurtosis |
|---|---|---|---|---|---|---|
| GRCS8 | -3.18644603 | 1 | 1.7291242 | 1.2705242 | 1.7964705 | 5.372929 |
| GRCS9 | -0.61378377 | 1 | 2.5030550 | 1.6267819 | 0.6678887 | 2.094293 |
| GRCS10 | -1.70844086 | 1 | 2.0509165 | 1.4468312 | 1.2264448 | 3.384691 |
| GRCS11 | -1.48706801 | 1 | 2.1547862 | 1.5294632 | 1.0848348 | 2.940572 |
| GRCS12 | -2.24236260 | 1 | 1.8411405 | 1.2806320 | 1.6467134 | 5.022449 |
| GRCS13 | -5.77221627 | 1 | 1.4643585 | 1.0576507 | 2.6371297 | 9.670295 |
| GRCS14 | -0.80735325 | 1 | 2.3645621 | 1.5582194 | 0.8034611 | 2.501572 |
| GRCS15 | -0.58697373 | 1 | 2.5539715 | 1.6838487 | 0.6895946 | 2.172498 |
| GRCS16 | -0.39066085 | 1 | 2.7026477 | 1.7406230 | 0.6450756 | 2.185271 |
| GRCS17 | -1.76406220 | 1 | 2.3503055 | 2.0422814 | 1.3082822 | 3.219497 |
| GRCS18 | -2.74077635 | 1 | 1.8289206 | 1.3639897 | 1.5668679 | 4.216822 |
| GRCS19 | -0.54890382 | 1 | 2.5784114 | 1.6937648 | 0.5977217 | 1.941036 |
| GRCS20 | -0.33281842 | 1 | 2.7026477 | 1.7205757 | 0.5329942 | 1.897831 |
| GRCS21 | -3.60192833 | 1 | 1.6415479 | 1.1560951 | 2.0054720 | 6.694470 |
| GRCS22 | -1.43565252 | 1 | 2.1812627 | 1.5537103 | 1.0887194 | 2.980026 |
| GRCS23 | -1.62888488 | 1 | 2.1446029 | 1.5732024 | 1.1739257 | 3.222479 |

Note. GRCS = Gambling Related Cognitions Scale; PDI = Peters Delusion Inventory; SBQ = Superstitious Belief Questionnaire; GCB = Generic Conspiracist Beliefs Scale.

**Table 3**

Kaiser-Meyer-Olkin and Bartlett-test

| Test | Value |
|---|---|
| KMO Overall | 0.942 |
| Bartlett Chi-square | 21,068.160 |
| Bartlett df | 3,403.000 |
| Bartlett p | 0.000 |

### Factor Extraction

**Table 4**

EFA factor eigenvalues and cumulative variance for the first 20 Factors

| Factor | Eigenvalue | CumulativeVar |
|---|---|---|
| F1 | 20.05 | 0.242 |
| F2 | 6.75 | 0.323 |
| F3 | 4.76 | 0.380 |
| F4 | 2.46 | 0.410 |
| F5 | 1.96 | 0.434 |
| F6 | 1.77 | 0.455 |
| F7 | 1.67 | 0.475 |
| F8 | 1.39 | 0.492 |
| F9 | 1.38 | 0.508 |
| F10 | 1.33 | 0.524 |
| F11 | 1.25 | 0.540 |

| Factor | Eigenvalue | CumulativeVar |
|---|---|---|
| F12 | 1.18 | 0.554 |
| F13 | 1.17 | 0.568 |
| F14 | 1.11 | 0.581 |
| F15 | 1.08 | 0.594 |
| F16 | 1.06 | 0.607 |
| F17 | 1.00 | 0.619 |
| F18 | 0.99 | 0.631 |
| F19 | 0.92 | 0.642 |
| F20 | 0.92 | 0.653 |

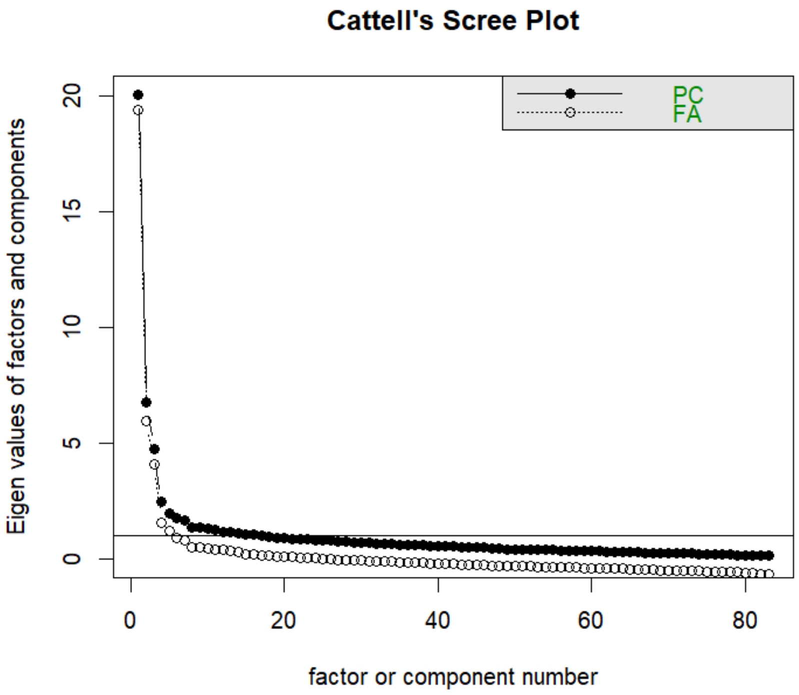


**Figure 1.** Cattell's Scree Plot

### Factor Loadings

**Table 3**

Factor Loadings for 3-Factor EFA

| Item | ML1 | ML2 | ML3 | h2 | u2 |
|---|---|---|---|---|---|
| PDI1 | 0.04 | 0.19 | 0.11 | 0.06963161 | 0.9303684 |
| PDI2 | -0.02 | 0.07 | 0.14 | 0.02846976 | 0.9715302 |
| PDI3 | 0.07 | 0.11 | 0.25 | 0.11849700 | 0.8815030 |
| PDI4 | 0.01 | 0.16 | 0.04 | 0.03287840 | 0.9671216 |
| PDI5 | 0.03 | -0.01 | 0.16 | 0.02866527 | 0.9713347 |
| PDI6 | 0.19 | 0.11 | 0.11 | 0.10417743 | 0.8958226 |
| PDI7 | 0.09 | 0.17 | 0.08 | 0.06878067 | 0.9312193 |
| PDI8 | 0.31 | -0.11 | 0.12 | 0.12549463 | 0.8745054 |
| PDI9 | 0.44 | -0.05 | 0.05 | 0.20559023 | 0.7944098 |
| PDI10 | 0.04 | 0.06 | 0.25 | 0.08405722 | 0.9159428 |
| PDI11 | 0.28 | -0.06 | 0.03 | 0.07824735 | 0.9217526 |
| PDI12 | 0.44 | -0.05 | 0.05 | 0.20210800 | 0.7978920 |
| PDI13 | 0.09 | 0.08 | 0.01 | 0.01932298 | 0.9806770 |
| PDI14 | -0.02 | 0.11 | 0.06 | 0.01660356 | 0.9833964 |
| PDI15 | -0.03 | 0.13 | 0.09 | 0.02830305 | 0.9716970 |
| PDI16 | 0.02 | 0.09 | 0.06 | 0.01687138 | 0.9831286 |
| PDI17 | -0.13 | 0.08 | 0.15 | 0.02524001 | 0.9747600 |

| Item | ML1 | ML2 | ML3 | h2 | u2 |
|---|---|---|---|---|---|
| PDI18 | 0.01 | 0.14 | 0.13 | 0.05190945 | 0.9480906 |
| PDI19 | 0.08 | 0.13 | 0.11 | 0.05799767 | 0.9420023 |
| PDI20 | 0.05 | 0.00 | 0.12 | 0.02505224 | 0.9749478 |
| GCB1 | -0.16 | 0.06 | 0.78 | 0.53050935 | 0.4694907 |
| GCB2 | -0.04 | 0.04 | 0.75 | 0.55614571 | 0.4438543 |
| GCB3 | -0.09 | 0.07 | 0.76 | 0.54212741 | 0.4578726 |
| GCB4 | -0.02 | -0.03 | 0.81 | 0.63614210 | 0.3638579 |
| GCB5 | -0.01 | -0.01 | 0.79 | 0.60862180 | 0.3913782 |
| GCB6 | 0.04 | -0.04 | 0.82 | 0.68341183 | 0.3165882 |
| GCB7 | 0.20 | -0.04 | 0.56 | 0.45052551 | 0.5494745 |
| GCB8 | 0.16 | -0.04 | 0.58 | 0.43285714 | 0.5671429 |
| GCB9 | 0.14 | 0.06 | 0.63 | 0.53292842 | 0.4670716 |
| GCB10 | 0.12 | 0.06 | 0.70 | 0.61700769 | 0.3829923 |
| GCB11 | 0.23 | -0.04 | 0.53 | 0.43792182 | 0.5620782 |
| GCB12 | 0.09 | 0.04 | 0.72 | 0.61285863 | 0.3871414 |
| GCB13 | 0.10 | -0.03 | 0.72 | 0.58707661 | 0.4129234 |
| GCB14 | -0.03 | 0.03 | 0.67 | 0.43669427 | 0.5633057 |
| GCB15 | -0.02 | -0.01 | 0.72 | 0.49885347 | 0.5011465 |
| SBQ1 | 0.81 | 0.03 | -0.12 | 0.58671806 | 0.4132819 |

| Item | ML1 | ML2 | ML3 | h2 | u2 |
|---|---|---|---|---|---|
| SBQ2 | 0.72 | 0.06 | 0.00 | 0.54566434 | 0.4543357 |
| SBQ3 | 0.82 | -0.03 | -0.04 | 0.61989768 | 0.3801023 |
| SBQ4 | 0.63 | 0.03 | 0.07 | 0.46851427 | 0.5314857 |
| SBQ5 | 0.73 | -0.03 | 0.10 | 0.60123599 | 0.3987640 |
| SBQ6 | 0.52 | 0.10 | -0.09 | 0.27261395 | 0.7273861 |
| SBQ7 | 0.76 | 0.01 | -0.03 | 0.55778617 | 0.4422138 |
| SBQ8 | 0.51 | -0.04 | 0.13 | 0.32868487 | 0.6713151 |
| SBQ9 | 0.71 | -0.04 | -0.01 | 0.47348307 | 0.5265169 |
| SBQ10 | 0.34 | 0.08 | -0.04 | 0.12842284 | 0.8715772 |
| SBQ11 | 0.75 | 0.08 | -0.07 | 0.56017672 | 0.4398233 |
| SBQ12 | 0.43 | -0.01 | 0.01 | 0.18904619 | 0.8109538 |
| SBQ13 | 0.50 | 0.04 | 0.17 | 0.38916914 | 0.6108309 |
| SBQ14 | 0.69 | -0.04 | 0.10 | 0.53695637 | 0.4630436 |
| SBQ15 | 0.68 | -0.09 | 0.07 | 0.47351473 | 0.5264853 |
| SBQ16 | 0.54 | 0.08 | 0.14 | 0.43757474 | 0.5624253 |
| SBQ17 | 0.50 | -0.02 | 0.15 | 0.33444889 | 0.6655511 |
| SBQ18 | 0.58 | -0.07 | 0.10 | 0.36982610 | 0.6301739 |
| SBQ19 | 0.75 | -0.03 | 0.09 | 0.61153258 | 0.3884674 |
| SBQ20 | 0.75 | 0.06 | 0.01 | 0.60215725 | 0.3978427 |

| Item | ML1 | ML2 | ML3 | h2 | u2 |
|---|---|---|---|---|---|
| SBQ21 | 0.66 | 0.12 | -0.03 | 0.48910951 | 0.5108905 |
| SBQ22 | 0.62 | 0.11 | 0.02 | 0.46683331 | 0.5331667 |
| SBQ23 | 0.66 | -0.01 | 0.06 | 0.46481254 | 0.5351875 |
| SBQ24 | 0.68 | -0.02 | 0.03 | 0.47042755 | 0.5295725 |
| SBQ25 | 0.65 | 0.00 | 0.06 | 0.46635778 | 0.5336422 |
| GRCS1 | -0.12 | 0.63 | 0.03 | 0.36968790 | 0.6303121 |
| GRCS2 | -0.02 | 0.55 | 0.01 | 0.30076934 | 0.6992307 |
| GRCS3 | 0.35 | 0.34 | 0.01 | 0.32863203 | 0.6713680 |
| GRCS4 | 0.15 | 0.57 | -0.03 | 0.39171589 | 0.6082841 |
| GRCS5 | 0.02 | 0.68 | -0.03 | 0.46136119 | 0.5386388 |
| GRCS6 | -0.01 | 0.62 | -0.05 | 0.36888985 | 0.6311101 |
| GRCS7 | -0.08 | 0.63 | 0.04 | 0.37648426 | 0.6235157 |
| GRCS8 | 0.28 | 0.51 | -0.04 | 0.41765626 | 0.5823437 |
| GRCS9 | 0.07 | 0.54 | 0.03 | 0.34413602 | 0.6558640 |
| GRCS10 | 0.06 | 0.69 | -0.06 | 0.48425462 | 0.5157454 |
| GRCS11 | 0.02 | 0.61 | -0.01 | 0.38096285 | 0.6190371 |
| GRCS12 | -0.07 | 0.64 | 0.12 | 0.42857660 | 0.5714234 |
| GRCS13 | 0.17 | 0.50 | 0.03 | 0.35429331 | 0.6457067 |
| GRCS14 | 0.17 | 0.61 | -0.05 | 0.44633271 | 0.5536673 |

| Item | ML1 | ML2 | ML3 | h2 | u2 |
|---|---|---|---|---|---|
| GRCS15 | -0.05 | 0.71 | 0.09 | 0.51224930 | 0.4877507 |
| GRCS16 | -0.11 | 0.57 | -0.04 | 0.28760642 | 0.7123936 |
| GRCS17 | -0.09 | 0.57 | 0.00 | 0.29251669 | 0.7074833 |
| GRCS18 | 0.21 | 0.54 | 0.00 | 0.41976181 | 0.5802382 |
| GRCS19 | -0.01 | 0.60 | 0.03 | 0.36565161 | 0.6343484 |
| GRCS20 | -0.04 | 0.68 | 0.06 | 0.47213799 | 0.5278620 |
| GRCS21 | -0.08 | 0.65 | 0.05 | 0.40253595 | 0.5974640 |
| GRCS22 | 0.03 | 0.58 | 0.07 | 0.38300474 | 0.6169953 |
| GRCS23 | 0.29 | 0.40 | 0.00 | 0.32467172 | 0.6753283 |

Note. GRCS = Gambling Related Cognitions Scale; PDI = Peters Delusion Inventory; SBQ = Superstitious Belief Questionnaire; GCB = Generic Conspiracist Beliefs Scale.

**Table 4**

Factor Loadings for 4-Factor EFA

| Item | ML1 | ML2 | ML3 | ML4 | h2 | u2 |
|---|---|---|---|---|---|---|
| PDI1 | 0.06 | 0.20 | 0.09 | 0.15 | 0.06963161 | 0.9303684 |
| PDI2 | -0.01 | 0.08 | 0.13 | 0.07 | 0.02846976 | 0.9715302 |
| PDI3 | 0.10 | 0.13 | 0.22 | 0.17 | 0.11849700 | 0.8815030 |
| PDI4 | 0.02 | 0.16 | 0.03 | 0.06 | 0.03287840 | 0.9671216 |
| PDI5 | 0.03 | -0.01 | 0.15 | 0.05 | 0.02866527 | 0.9713347 |
| PDI6 | 0.21 | 0.12 | 0.10 | 0.12 | 0.10417743 | 0.8958226 |

| Item | ML1 | ML2 | ML3 | ML4 | h2 | u2 |
|---|---|---|---|---|---|---|
| PDI7 | 0.13 | 0.19 | 0.04 | 0.22 | 0.06878067 | 0.9312193 |
| PDI8 | 0.33 | -0.09 | 0.09 | 0.19 | 0.12549463 | 0.8745054 |
| PDI9 | 0.43 | -0.04 | 0.06 | 0.00 | 0.20559023 | 0.7944098 |
| PDI10 | 0.07 | 0.07 | 0.22 | 0.18 | 0.08405722 | 0.9159428 |
| PDI11 | 0.29 | -0.05 | 0.02 | 0.08 | 0.07824735 | 0.9217526 |
| PDI12 | 0.43 | -0.04 | 0.06 | -0.01 | 0.20210800 | 0.7978920 |
| PDI13 | 0.11 | 0.09 | -0.01 | 0.12 | 0.01932298 | 0.9806770 |
| PDI14 | -0.01 | 0.11 | 0.05 | 0.07 | 0.01660356 | 0.9833964 |
| PDI15 | 0.00 | 0.14 | 0.07 | 0.15 | 0.02830305 | 0.9716970 |
| PDI16 | 0.04 | 0.10 | 0.03 | 0.14 | 0.01687138 | 0.9831286 |
| PDI17 | -0.11 | 0.08 | 0.13 | 0.09 | 0.02524001 | 0.9747600 |
| PDI18 | 0.03 | 0.15 | 0.12 | 0.06 | 0.05190945 | 0.9480906 |
| PDI19 | 0.10 | 0.13 | 0.09 | 0.12 | 0.05799767 | 0.9420023 |
| PDI20 | 0.06 | 0.00 | 0.12 | 0.04 | 0.02505224 | 0.9749478 |
| GCB1 | -0.14 | 0.06 | 0.76 | 0.11 | 0.53050935 | 0.4694907 |
| GCB2 | -0.06 | 0.03 | 0.77 | -0.08 | 0.55614571 | 0.4438543 |
| GCB3 | -0.07 | 0.08 | 0.74 | 0.14 | 0.54212741 | 0.4578726 |
| GCB4 | -0.03 | -0.03 | 0.82 | -0.04 | 0.63614210 | 0.3638579 |
| GCB5 | -0.02 | -0.01 | 0.79 | -0.02 | 0.60862180 | 0.3913782 |

| Item | ML1 | ML2 | ML3 | ML4 | h2 | u2 |
|---|---|---|---|---|---|---|
| GCB6 | 0.03 | -0.04 | 0.82 | 0.00 | 0.68341183 | 0.3165882 |
| GCB7 | 0.15 | -0.05 | 0.61 | -0.23 | 0.45052551 | 0.5494745 |
| GCB8 | 0.12 | -0.04 | 0.61 | -0.13 | 0.43285714 | 0.5671429 |
| GCB9 | 0.09 | 0.05 | 0.68 | -0.21 | 0.53292842 | 0.4670716 |
| GCB10 | 0.10 | 0.06 | 0.71 | -0.07 | 0.61700769 | 0.3829923 |
| GCB11 | 0.21 | -0.04 | 0.54 | -0.06 | 0.43792182 | 0.5620782 |
| GCB12 | 0.09 | 0.04 | 0.72 | 0.00 | 0.61285863 | 0.3871414 |
| GCB13 | 0.11 | -0.02 | 0.71 | 0.07 | 0.58707661 | 0.4129234 |
| GCB14 | -0.01 | 0.04 | 0.65 | 0.13 | 0.43669427 | 0.5633057 |
| GCB15 | 0.02 | 0.01 | 0.68 | 0.24 | 0.49885347 | 0.5011465 |
| SBQ1 | 0.77 | 0.03 | -0.08 | -0.19 | 0.58671806 | 0.4132819 |
| SBQ2 | 0.66 | 0.06 | 0.06 | -0.33 | 0.54566434 | 0.4543357 |
| SBQ3 | 0.77 | -0.03 | 0.01 | -0.27 | 0.61989768 | 0.3801023 |
| SBQ4 | 0.67 | 0.06 | 0.04 | 0.24 | 0.46851427 | 0.5314857 |
| SBQ5 | 0.74 | -0.01 | 0.09 | 0.14 | 0.60123599 | 0.3987640 |
| SBQ6 | 0.47 | 0.10 | -0.05 | -0.20 | 0.27261395 | 0.7273861 |
| SBQ7 | 0.72 | 0.02 | 0.01 | -0.13 | 0.55778617 | 0.4422138 |
| SBQ8 | 0.53 | -0.02 | 0.12 | 0.17 | 0.32868487 | 0.6713151 |
| SBQ9 | 0.75 | -0.01 | -0.05 | 0.25 | 0.47348307 | 0.5265169 |

| Item | ML1 | ML2 | ML3 | ML4 | h2 | u2 |
|---|---|---|---|---|---|---|
| SBQ10 | 0.36 | 0.09 | -0.05 | 0.11 | 0.12842284 | 0.8715772 |
| SBQ11 | 0.72 | 0.09 | -0.03 | -0.11 | 0.56017672 | 0.4398233 |
| SBQ12 | 0.44 | 0.00 | 0.01 | 0.08 | 0.18904619 | 0.8109538 |
| SBQ13 | 0.48 | 0.05 | 0.19 | -0.02 | 0.38916914 | 0.6108309 |
| SBQ14 | 0.66 | -0.03 | 0.14 | -0.11 | 0.53695637 | 0.4630436 |
| SBQ15 | 0.68 | -0.06 | 0.06 | 0.09 | 0.47351473 | 0.5264853 |
| SBQ16 | 0.55 | 0.10 | 0.14 | 0.11 | 0.43757474 | 0.5624253 |
| SBQ17 | 0.54 | 0.01 | 0.11 | 0.30 | 0.33444889 | 0.6655511 |
| SBQ18 | 0.60 | -0.04 | 0.07 | 0.22 | 0.36982610 | 0.6301739 |
| SBQ19 | 0.72 | -0.01 | 0.11 | -0.03 | 0.61153258 | 0.3884674 |
| SBQ20 | 0.70 | 0.07 | 0.05 | -0.19 | 0.60215725 | 0.3978427 |
| SBQ21 | 0.63 | 0.13 | 0.00 | -0.08 | 0.48910951 | 0.5108905 |
| SBQ22 | 0.63 | 0.13 | 0.02 | 0.08 | 0.46683331 | 0.5331667 |
| SBQ23 | 0.67 | 0.01 | 0.05 | 0.13 | 0.46481254 | 0.5351875 |
| SBQ24 | 0.71 | 0.01 | 0.01 | 0.20 | 0.47042755 | 0.5295725 |
| SBQ25 | 0.64 | 0.02 | 0.07 | 0.01 | 0.46635778 | 0.5336422 |
| GRCS1 | -0.11 | 0.63 | 0.02 | 0.08 | 0.36968790 | 0.6303121 |
| GRCS2 | -0.06 | 0.54 | 0.04 | -0.20 | 0.30076934 | 0.6992307 |
| GRCS3 | 0.33 | 0.35 | 0.03 | -0.08 | 0.32863203 | 0.6713680 |

| Item | ML1 | ML2 | ML3 | ML4 | h2 | u2 |
|---|---|---|---|---|---|---|
| GRCS4 | 0.13 | 0.56 | -0.01 | -0.14 | 0.39171589 | 0.6082841 |
| GRCS5 | 0.05 | 0.69 | -0.06 | 0.17 | 0.46136119 | 0.5386388 |
| GRCS6 | 0.00 | 0.63 | -0.06 | 0.09 | 0.36888985 | 0.6311101 |
| GRCS7 | -0.10 | 0.62 | 0.06 | -0.13 | 0.37648426 | 0.6235157 |
| GRCS8 | 0.25 | 0.51 | -0.01 | -0.14 | 0.41765626 | 0.5823437 |
| GRCS9 | 0.09 | 0.55 | 0.02 | 0.11 | 0.34413602 | 0.6558640 |
| GRCS10 | 0.06 | 0.69 | -0.07 | 0.01 | 0.48425462 | 0.5157454 |
| GRCS11 | 0.03 | 0.61 | -0.01 | 0.02 | 0.38096285 | 0.6190371 |
| GRCS12 | -0.08 | 0.63 | 0.13 | -0.04 | 0.42857660 | 0.5714234 |
| GRCS13 | 0.13 | 0.49 | 0.07 | -0.23 | 0.35429331 | 0.6457067 |
| GRCS14 | 0.17 | 0.61 | -0.05 | -0.01 | 0.44633271 | 0.5536673 |
| GRCS15 | -0.04 | 0.71 | 0.08 | 0.07 | 0.51224930 | 0.4877507 |
| GRCS16 | -0.09 | 0.57 | -0.06 | 0.10 | 0.28760642 | 0.7123936 |
| GRCS17 | -0.10 | 0.56 | 0.01 | -0.09 | 0.29251669 | 0.7074833 |
| GRCS18 | 0.19 | 0.54 | 0.02 | -0.11 | 0.41976181 | 0.5802382 |
| GRCS19 | 0.01 | 0.60 | 0.01 | 0.06 | 0.36565161 | 0.6343484 |
| GRCS20 | -0.01 | 0.69 | 0.04 | 0.12 | 0.47213799 | 0.5278620 |
| GRCS21 | -0.11 | 0.63 | 0.08 | -0.21 | 0.40253595 | 0.5974640 |
| GRCS22 | 0.05 | 0.59 | 0.06 | 0.10 | 0.38300474 | 0.6169953 |

| Item | ML1 | ML2 | ML3 | ML4 | h2 | u2 |
|---|---|---|---|---|---|---|
| GRCS23 | 0.26 | 0.40 | 0.02 | -0.12 | 0.32467172 | 0.6753283 |

Note. GRCS = Gambling Related Cognitions Scale; PDI = Peters Delusion Inventory; SBQ = Superstitious Belief Questionnaire; GCB = Generic Conspiracist Beliefs Scale.

**Table 5**

Factor Loadings bifactor EFA

| Item | ML1 | ML2 | ML3 | ML4 | h2 | u2 |
|---|---|---|---|---|---|---|
| PDI1 | 0.15 | 0.19 | 0.12 | 0.14 | 0.09 | 0.91 |
| PDI2 | 0.09 | 0.08 | 0.13 | 0.05 | 0.03 | 0.97 |
| PDI3 | 0.24 | 0.13 | 0.22 | 0.17 | 0.15 | 0.85 |
| PDI4 | 0.09 | 0.15 | 0.04 | 0.06 | 0.04 | 0.96 |
| PDI5 | 0.11 | 0.00 | 0.13 | 0.04 | 0.03 | 0.97 |
| PDI6 | 0.27 | 0.11 | 0.08 | 0.17 | 0.12 | 0.88 |
| PDI7 | 0.15 | 0.20 | 0.10 | 0.23 | 0.12 | 0.88 |
| PDI8 | 0.25 | -0.08 | 0.10 | 0.29 | 0.16 | 0.84 |
| PDI9 | 0.41 | -0.07 | -0.02 | 0.17 | 0.20 | 0.80 |
| PDI10 | 0.18 | 0.08 | 0.22 | 0.16 | 0.11 | 0.89 |
| PDI11 | 0.22 | -0.05 | 0.00 | 0.18 | 0.09 | 0.91 |
| PDI12 | 0.41 | -0.07 | -0.03 | 0.17 | 0.20 | 0.80 |
| PDI13 | 0.09 | 0.09 | 0.03 | 0.14 | 0.03 | 0.97 |
| PDI14 | 0.05 | 0.11 | 0.07 | 0.04 | 0.02 | 0.98 |

| Item | ML1 | ML2 | ML3 | ML4 | h2 | u2 |
|---|---|---|---|---|---|---|
| PDI15 | 0.06 | 0.15 | 0.11 | 0.11 | 0.05 | 0.95 |
| PDI16 | 0.06 | 0.11 | 0.07 | 0.12 | 0.04 | 0.96 |
| PDI17 | 0.00 | 0.09 | 0.15 | 0.02 | 0.03 | 0.97 |
| PDI18 | 0.15 | 0.14 | 0.11 | 0.05 | 0.06 | 0.94 |
| PDI19 | 0.17 | 0.13 | 0.10 | 0.13 | 0.07 | 0.93 |
| PDI20 | 0.12 | 0.00 | 0.09 | 0.05 | 0.03 | 0.97 |
| GCB1 | 0.38 | 0.06 | 0.62 | 0.00 | 0.54 | 0.46 |
| GCB2 | 0.52 | 0.00 | 0.54 | -0.11 | 0.57 | 0.43 |
| GCB3 | 0.43 | 0.08 | 0.60 | 0.06 | 0.55 | 0.45 |
| GCB4 | 0.54 | -0.05 | 0.59 | -0.07 | 0.64 | 0.36 |
| GCB5 | 0.53 | -0.03 | 0.57 | -0.05 | 0.61 | 0.39 |
| GCB6 | 0.57 | -0.06 | 0.59 | -0.01 | 0.68 | 0.32 |
| GCB7 | 0.61 | -0.11 | 0.33 | -0.14 | 0.51 | 0.49 |
| GCB8 | 0.56 | -0.09 | 0.37 | -0.08 | 0.46 | 0.54 |
| GCB9 | 0.64 | -0.01 | 0.39 | -0.16 | 0.59 | 0.41 |
| GCB10 | 0.63 | 0.02 | 0.47 | -0.04 | 0.62 | 0.38 |
| GCB11 | 0.57 | -0.08 | 0.34 | 0.02 | 0.44 | 0.56 |
| GCB12 | 0.59 | 0.01 | 0.51 | 0.01 | 0.61 | 0.39 |
| GCB13 | 0.55 | -0.04 | 0.53 | 0.07 | 0.59 | 0.41 |

| Item | ML1 | ML2 | ML3 | ML4 | h2 | u2 |
|---|---|---|---|---|---|---|
| GCB14 | 0.40 | 0.04 | 0.53 | 0.08 | 0.45 | 0.55 |
| GCB15 | 0.40 | 0.03 | 0.60 | 0.18 | 0.55 | 0.45 |
| SBQ1 | 0.72 | -0.06 | -0.26 | 0.16 | 0.62 | 0.38 |
| SBQ2 | 0.78 | -0.06 | -0.21 | -0.01 | 0.65 | 0.35 |
| SBQ3 | 0.78 | -0.13 | -0.23 | 0.09 | 0.69 | 0.31 |
| SBQ4 | 0.57 | 0.04 | 0.02 | 0.46 | 0.54 | 0.46 |
| SBQ5 | 0.68 | -0.05 | 0.01 | 0.41 | 0.63 | 0.37 |
| SBQ6 | 0.51 | 0.02 | -0.20 | 0.03 | 0.30 | 0.70 |
| SBQ7 | 0.71 | -0.07 | -0.17 | 0.18 | 0.57 | 0.43 |
| SBQ8 | 0.48 | -0.04 | 0.07 | 0.35 | 0.36 | 0.64 |
| SBQ9 | 0.54 | -0.03 | -0.05 | 0.51 | 0.56 | 0.44 |
| SBQ10 | 0.29 | 0.07 | -0.05 | 0.23 | 0.14 | 0.86 |
| SBQ11 | 0.70 | 0.00 | -0.19 | 0.19 | 0.56 | 0.44 |
| SBQ12 | 0.37 | -0.02 | -0.03 | 0.24 | 0.20 | 0.80 |
| SBQ13 | 0.60 | 0.00 | 0.05 | 0.17 | 0.39 | 0.61 |
| SBQ14 | 0.71 | -0.10 | -0.05 | 0.17 | 0.54 | 0.46 |
| SBQ15 | 0.59 | -0.10 | -0.02 | 0.35 | 0.49 | 0.51 |
| SBQ16 | 0.59 | 0.06 | 0.05 | 0.31 | 0.45 | 0.55 |
| SBQ17 | 0.46 | 0.01 | 0.12 | 0.46 | 0.44 | 0.56 |

| Item | ML1 | ML2 | ML3 | ML4 | h2 | u2 |
|---|---|---|---|---|---|---|
| SBQ18 | 0.49 | -0.05 | 0.05 | 0.42 | 0.42 | 0.58 |
| SBQ19 | 0.73 | -0.08 | -0.05 | 0.26 | 0.61 | 0.39 |
| SBQ20 | 0.77 | -0.03 | -0.15 | 0.12 | 0.63 | 0.37 |
| SBQ21 | 0.66 | 0.05 | -0.14 | 0.18 | 0.49 | 0.51 |
| SBQ22 | 0.61 | 0.08 | -0.06 | 0.31 | 0.48 | 0.52 |
| SBQ23 | 0.59 | -0.02 | -0.02 | 0.37 | 0.49 | 0.51 |
| SBQ24 | 0.57 | -0.02 | -0.03 | 0.44 | 0.52 | 0.48 |
| SBQ25 | 0.63 | -0.03 | -0.05 | 0.26 | 0.47 | 0.53 |
| GRCS1 | 0.18 | 0.58 | 0.05 | 0.00 | 0.38 | 0.62 |
| GRCS2 | 0.29 | 0.45 | -0.06 | -0.21 | 0.34 | 0.66 |
| GRCS3 | 0.50 | 0.27 | -0.07 | 0.05 | 0.33 | 0.67 |
| GRCS4 | 0.42 | 0.47 | -0.10 | -0.08 | 0.41 | 0.59 |
| GRCS5 | 0.26 | 0.64 | 0.01 | 0.14 | 0.50 | 0.50 |
| GRCS6 | 0.21 | 0.57 | -0.02 | 0.05 | 0.38 | 0.62 |
| GRCS7 | 0.28 | 0.53 | -0.01 | -0.17 | 0.39 | 0.61 |
| GRCS8 | 0.50 | 0.41 | -0.11 | -0.03 | 0.43 | 0.57 |
| GRCS9 | 0.31 | 0.50 | 0.03 | 0.10 | 0.36 | 0.64 |
| GRCS10 | 0.32 | 0.61 | -0.07 | 0.01 | 0.49 | 0.51 |
| GRCS11 | 0.28 | 0.55 | -0.02 | 0.00 | 0.38 | 0.62 |

| Item | ML1 | ML2 | ML3 | ML4 | h2 | u2 |
|---|---|---|---|---|---|---|
| GRCS12 | 0.32 | 0.56 | 0.08 | -0.09 | 0.43 | 0.57 |
| GRCS13 | 0.47 | 0.39 | -0.08 | -0.16 | 0.40 | 0.60 |
| GRCS14 | 0.40 | 0.53 | -0.08 | 0.04 | 0.44 | 0.56 |
| GRCS15 | 0.31 | 0.64 | 0.08 | 0.01 | 0.52 | 0.48 |
| GRCS16 | 0.10 | 0.54 | 0.00 | 0.03 | 0.30 | 0.70 |
| GRCS17 | 0.20 | 0.49 | -0.02 | -0.13 | 0.30 | 0.70 |
| GRCS18 | 0.47 | 0.45 | -0.07 | -0.04 | 0.43 | 0.57 |
| GRCS19 | 0.27 | 0.55 | 0.02 | 0.03 | 0.37 | 0.63 |
| GRCS20 | 0.28 | 0.63 | 0.06 | 0.06 | 0.49 | 0.51 |
| GRCS21 | 0.31 | 0.54 | -0.03 | -0.24 | 0.45 | 0.55 |
| GRCS22 | 0.31 | 0.54 | 0.07 | 0.07 | 0.40 | 0.60 |
| GRCS23 | 0.48 | 0.32 | -0.08 | 0.00 | 0.33 | 0.67 |

Note. GRCS = Gambling Related Cognitions Scale; PDI = Peters Delusion Inventory; SBQ = Superstitious Belief Questionnaire; GCB = Generic Conspiracist Beliefs Scale.

**Table 6**

Factor Loadings orthogonal CFA

| Factor | Item | Estimate | SE | Z | p | Std.Loading | CI.Lower | CI.Upper |
|---|---|---|---|---|---|---|---|---|
| SF | SBQ1 | 0.753 | 0.039 | 19.469 | 0 | 0.754 | 0.677 | 0.829 |
| SF | SBQ2 | 0.727 | 0.039 | 18.540 | 0 | 0.728 | 0.650 | 0.804 |
| SF | SBQ3 | 0.781 | 0.038 | 20.503 | 0 | 0.781 | 0.706 | 0.855 |

| Factor | Item | Estimate | SE | Z | p | Std.Loading | CI.Lower | CI.Upper |
|---|---|---|---|---|---|---|---|---|
| SF | SBQ4 | 0.691 | 0.040 | 17.288 | 0 | 0.691 | 0.612 | 0.769 |
| SF | SBQ5 | 0.774 | 0.038 | 20.238 | 0 | 0.774 | 0.699 | 0.849 |
| SF | SBQ6 | 0.495 | 0.043 | 11.510 | 0 | 0.496 | 0.411 | 0.579 |
| SF | SBQ7 | 0.744 | 0.039 | 19.144 | 0 | 0.745 | 0.668 | 0.820 |
| SF | SBQ8 | 0.572 | 0.042 | 13.643 | 0 | 0.573 | 0.490 | 0.654 |
| SF | SBQ9 | 0.687 | 0.040 | 17.165 | 0 | 0.688 | 0.608 | 0.765 |
| SF | SBQ10 | 0.349 | 0.045 | 7.837 | 0 | 0.350 | 0.262 | 0.437 |
| SF | SBQ11 | 0.742 | 0.039 | 19.069 | 0 | 0.743 | 0.666 | 0.818 |
| SF | SBQ12 | 0.448 | 0.044 | 10.292 | 0 | 0.449 | 0.363 | 0.534 |
| SF | SBQ13 | 0.612 | 0.041 | 14.811 | 0 | 0.613 | 0.531 | 0.693 |
| SF | SBQ14 | 0.725 | 0.039 | 18.476 | 0 | 0.726 | 0.648 | 0.802 |
| SF | SBQ15 | 0.684 | 0.040 | 17.077 | 0 | 0.685 | 0.606 | 0.763 |
| SF | SBQ16 | 0.651 | 0.041 | 16.005 | 0 | 0.652 | 0.571 | 0.731 |
| SF | SBQ17 | 0.579 | 0.042 | 13.831 | 0 | 0.579 | 0.497 | 0.661 |
| SF | SBQ18 | 0.600 | 0.042 | 14.448 | 0 | 0.600 | 0.518 | 0.681 |
| SF | SBQ19 | 0.779 | 0.038 | 20.436 | 0 | 0.780 | 0.704 | 0.854 |
| SF | SBQ20 | 0.770 | 0.038 | 20.086 | 0 | 0.770 | 0.695 | 0.845 |
| SF | SBQ21 | 0.683 | 0.040 | 17.027 | 0 | 0.683 | 0.604 | 0.761 |
| SF | SBQ22 | 0.678 | 0.040 | 16.878 | 0 | 0.679 | 0.599 | 0.757 |

| Factor | Item | Estimate | SE | Z | p | Std.Loading | CI.Lower | CI.Upper |
|---|---|---|---|---|---|---|---|---|
| SF | SBQ23 | 0.691 | 0.040 | 17.303 | 0 | 0.692 | 0.613 | 0.769 |
| SF | SBQ24 | 0.689 | 0.040 | 17.244 | 0 | 0.690 | 0.611 | 0.768 |
| SF | SBQ25 | 0.686 | 0.040 | 17.125 | 0 | 0.686 | 0.607 | 0.764 |
| SF | PDI9 | 0.443 | 0.044 | 10.149 | 0 | 0.443 | 0.357 | 0.528 |
| SF | PDI12 | 0.438 | 0.044 | 10.023 | 0 | 0.438 | 0.352 | 0.524 |
| GF | GRCS1 | 0.585 | 0.042 | 13.848 | 0 | 0.586 | 0.502 | 0.668 |
| GF | GRCS2 | 0.539 | 0.043 | 12.551 | 0 | 0.540 | 0.455 | 0.623 |
| GF | GRCS3 | 0.492 | 0.044 | 11.290 | 0 | 0.493 | 0.407 | 0.578 |
| GF | GRCS4 | 0.632 | 0.041 | 15.233 | 0 | 0.632 | 0.550 | 0.713 |
| GF | GRCS5 | 0.683 | 0.041 | 16.854 | 0 | 0.684 | 0.604 | 0.763 |
| GF | GRCS6 | 0.590 | 0.042 | 13.990 | 0 | 0.591 | 0.507 | 0.673 |
| GF | GRCS7 | 0.595 | 0.042 | 14.151 | 0 | 0.596 | 0.513 | 0.678 |
| GF | GRCS8 | 0.613 | 0.042 | 14.678 | 0 | 0.614 | 0.531 | 0.695 |
| GF | GRCS9 | 0.592 | 0.042 | 14.039 | 0 | 0.592 | 0.509 | 0.674 |
| GF | GRCS10 | 0.696 | 0.040 | 17.291 | 0 | 0.697 | 0.617 | 0.775 |
| GF | GRCS11 | 0.610 | 0.042 | 14.573 | 0 | 0.610 | 0.528 | 0.692 |
| GF | GRCS12 | 0.641 | 0.041 | 15.512 | 0 | 0.641 | 0.560 | 0.722 |
| GF | GRCS13 | 0.581 | 0.042 | 13.737 | 0 | 0.582 | 0.498 | 0.664 |
| GF | GRCS14 | 0.662 | 0.041 | 16.185 | 0 | 0.663 | 0.582 | 0.743 |

| Factor | Item | Estimate | SE | Z | p | Std.Loading | CI.Lower | CI.Upper |
|---|---|---|---|---|---|---|---|---|
| GF | GRCS15 | 0.709 | 0.040 | 17.715 | 0 | 0.710 | 0.630 | 0.787 |
| GF | GRCS16 | 0.500 | 0.043 | 11.499 | 0 | 0.501 | 0.415 | 0.585 |
| GF | GRCS17 | 0.517 | 0.043 | 11.966 | 0 | 0.518 | 0.433 | 0.602 |
| GF | GRCS18 | 0.632 | 0.041 | 15.232 | 0 | 0.632 | 0.550 | 0.713 |
| GF | GRCS19 | 0.602 | 0.042 | 14.335 | 0 | 0.602 | 0.519 | 0.684 |
| GF | GRCS20 | 0.684 | 0.041 | 16.878 | 0 | 0.685 | 0.604 | 0.763 |
| GF | GRCS21 | 0.618 | 0.042 | 14.824 | 0 | 0.619 | 0.536 | 0.700 |
| GF | GRCS22 | 0.619 | 0.042 | 14.846 | 0 | 0.620 | 0.537 | 0.701 |
| GF | GRCS23 | 0.526 | 0.043 | 12.195 | 0 | 0.526 | 0.441 | 0.610 |
| CF | GCB1 | 0.698 | 0.040 | 17.468 | 0 | 0.699 | 0.620 | 0.777 |
| CF | GCB2 | 0.745 | 0.039 | 19.106 | 0 | 0.746 | 0.669 | 0.822 |
| CF | GCB3 | 0.715 | 0.040 | 18.045 | 0 | 0.716 | 0.637 | 0.793 |
| CF | GCB4 | 0.797 | 0.038 | 21.063 | 0 | 0.798 | 0.723 | 0.871 |
| CF | GCB5 | 0.779 | 0.038 | 20.367 | 0 | 0.780 | 0.704 | 0.854 |
| CF | GCB6 | 0.827 | 0.037 | 22.286 | 0 | 0.828 | 0.755 | 0.900 |
| CF | GCB7 | 0.669 | 0.041 | 16.495 | 0 | 0.669 | 0.589 | 0.748 |
| CF | GCB8 | 0.656 | 0.041 | 16.104 | 0 | 0.657 | 0.576 | 0.736 |
| CF | GCB9 | 0.730 | 0.039 | 18.574 | 0 | 0.731 | 0.653 | 0.807 |
| CF | GCB10 | 0.783 | 0.038 | 20.508 | 0 | 0.784 | 0.708 | 0.858 |

| Factor | Item | Estimate | SE | Z | p | Std.Loading | CI.Lower | CI.Upper |
|---|---|---|---|---|---|---|---|---|
| CF | GCB11 | 0.640 | 0.041 | 15.594 | 0 | 0.641 | 0.560 | 0.720 |
| CF | GCB12 | 0.780 | 0.038 | 20.378 | 0 | 0.780 | 0.705 | 0.854 |
| CF | GCB13 | 0.764 | 0.039 | 19.785 | 0 | 0.765 | 0.688 | 0.839 |
| CF | GCB14 | 0.651 | 0.041 | 15.941 | 0 | 0.652 | 0.571 | 0.731 |
| CF | GCB15 | 0.692 | 0.040 | 17.252 | 0 | 0.692 | 0.613 | 0.770 |

Note. SF = Superstition Factor; GF = Gambling Factor; CF = Conspiracy Factor; GRCS = Gambling Related Cognitions Scale; PDI = Peters Delusion Inventory; SBQ = Superstitious Belief Questionnaire; GCB = Generic Conspiracist Beliefs Scale.

**Table 7**

Factor Loadings hierarchical CFA

| Factor | Item | Estimate | SE | Z | p | Std.Loading | CI.Lower | CI.Upper |
|---|---|---|---|---|---|---|---|---|
| SF | SBQ1 | 0.389 | 0.061 | 6.336 | 0 | 0.750 | 0.269 | 0.509 |
| SF | SBQ2 | 0.378 | 0.060 | 6.307 | 0 | 0.728 | 0.260 | 0.495 |
| SF | SBQ3 | 0.404 | 0.063 | 6.371 | 0 | 0.779 | 0.280 | 0.528 |
| SF | SBQ4 | 0.359 | 0.057 | 6.254 | 0 | 0.692 | 0.246 | 0.471 |
| SF | SBQ5 | 0.402 | 0.063 | 6.367 | 0 | 0.775 | 0.278 | 0.526 |
| SF | SBQ6 | 0.257 | 0.044 | 5.793 | 0 | 0.495 | 0.170 | 0.343 |
| SF | SBQ7 | 0.385 | 0.061 | 6.327 | 0 | 0.743 | 0.266 | 0.505 |
| SF | SBQ8 | 0.298 | 0.049 | 6.023 | 0 | 0.574 | 0.201 | 0.395 |
| SF | SBQ9 | 0.355 | 0.057 | 6.243 | 0 | 0.685 | 0.244 | 0.467 |

| Factor | Item | Estimate | SE | Z | p | Std.Loading | CI.Lower | CI.Upper |
|---|---|---|---|---|---|---|---|---|
| SF | SBQ10 | 0.181 | 0.036 | 5.095 | 0 | 0.349 | 0.111 | 0.251 |
| SF | SBQ11 | 0.384 | 0.061 | 6.325 | 0 | 0.741 | 0.265 | 0.504 |
| SF | SBQ12 | 0.232 | 0.041 | 5.614 | 0 | 0.448 | 0.151 | 0.313 |
| SF | SBQ13 | 0.320 | 0.052 | 6.119 | 0 | 0.617 | 0.217 | 0.422 |
| SF | SBQ14 | 0.377 | 0.060 | 6.306 | 0 | 0.727 | 0.260 | 0.494 |
| SF | SBQ15 | 0.355 | 0.057 | 6.241 | 0 | 0.684 | 0.243 | 0.466 |
| SF | SBQ16 | 0.340 | 0.055 | 6.194 | 0 | 0.656 | 0.232 | 0.448 |
| SF | SBQ17 | 0.302 | 0.050 | 6.040 | 0 | 0.581 | 0.204 | 0.399 |
| SF | SBQ18 | 0.312 | 0.051 | 6.084 | 0 | 0.601 | 0.211 | 0.412 |
| SF | SBQ19 | 0.405 | 0.063 | 6.373 | 0 | 0.780 | 0.280 | 0.529 |
| SF | SBQ20 | 0.400 | 0.063 | 6.362 | 0 | 0.771 | 0.277 | 0.523 |
| SF | SBQ21 | 0.355 | 0.057 | 6.242 | 0 | 0.684 | 0.243 | 0.466 |
| SF | SBQ22 | 0.353 | 0.057 | 6.235 | 0 | 0.680 | 0.242 | 0.464 |
| SF | SBQ23 | 0.359 | 0.057 | 6.253 | 0 | 0.692 | 0.246 | 0.471 |
| SF | SBQ24 | 0.357 | 0.057 | 6.249 | 0 | 0.689 | 0.245 | 0.469 |
| SF | SBQ25 | 0.356 | 0.057 | 6.246 | 0 | 0.687 | 0.244 | 0.468 |
| SF | PDI9 | 0.230 | 0.041 | 5.596 | 0 | 0.443 | 0.149 | 0.310 |
| SF | PDI12 | 0.227 | 0.041 | 5.573 | 0 | 0.438 | 0.147 | 0.307 |
| GF | GRCS1 | 0.497 | 0.038 | 13.159 | 0 | 0.580 | 0.423 | 0.571 |

| Factor | Item | Estimate | SE | Z | p | Std.Loading | CI.Lower | CI.Upper |
|---|---|---|---|---|---|---|---|---|
| GF | GRCS2 | 0.461 | 0.038 | 12.088 | 0 | 0.537 | 0.386 | 0.535 |
| GF | GRCS3 | 0.432 | 0.038 | 11.265 | 0 | 0.504 | 0.357 | 0.507 |
| GF | GRCS4 | 0.545 | 0.037 | 14.600 | 0 | 0.635 | 0.472 | 0.618 |
| GF | GRCS5 | 0.584 | 0.037 | 15.838 | 0 | 0.681 | 0.512 | 0.657 |
| GF | GRCS6 | 0.503 | 0.038 | 13.333 | 0 | 0.587 | 0.429 | 0.577 |
| GF | GRCS7 | 0.508 | 0.038 | 13.479 | 0 | 0.592 | 0.434 | 0.582 |
| GF | GRCS8 | 0.533 | 0.037 | 14.246 | 0 | 0.622 | 0.460 | 0.607 |
| GF | GRCS9 | 0.510 | 0.038 | 13.528 | 0 | 0.594 | 0.436 | 0.583 |
| GF | GRCS10 | 0.596 | 0.037 | 16.217 | 0 | 0.695 | 0.524 | 0.668 |
| GF | GRCS11 | 0.522 | 0.038 | 13.893 | 0 | 0.608 | 0.448 | 0.595 |
| GF | GRCS12 | 0.548 | 0.037 | 14.710 | 0 | 0.639 | 0.475 | 0.622 |
| GF | GRCS13 | 0.504 | 0.038 | 13.358 | 0 | 0.588 | 0.430 | 0.578 |
| GF | GRCS14 | 0.571 | 0.037 | 15.410 | 0 | 0.666 | 0.498 | 0.644 |
| GF | GRCS15 | 0.607 | 0.037 | 16.560 | 0 | 0.707 | 0.535 | 0.679 |
| GF | GRCS16 | 0.423 | 0.038 | 11.017 | 0 | 0.494 | 0.348 | 0.499 |
| GF | GRCS17 | 0.440 | 0.038 | 11.485 | 0 | 0.513 | 0.365 | 0.515 |
| GF | GRCS18 | 0.548 | 0.037 | 14.692 | 0 | 0.639 | 0.475 | 0.621 |
| GF | GRCS19 | 0.515 | 0.038 | 13.695 | 0 | 0.601 | 0.441 | 0.589 |
| GF | GRCS20 | 0.585 | 0.037 | 15.860 | 0 | 0.682 | 0.513 | 0.657 |

| Factor | Item | Estimate | SE | Z | p | Std.Loading | CI.Lower | CI.Upper |
|---|---|---|---|---|---|---|---|---|
| GF | GRCS21 | 0.527 | 0.038 | 14.063 | 0 | 0.615 | 0.454 | 0.601 |
| GF | GRCS22 | 0.533 | 0.037 | 14.222 | 0 | 0.621 | 0.459 | 0.606 |
| GF | GRCS23 | 0.459 | 0.038 | 12.042 | 0 | 0.535 | 0.384 | 0.534 |
| CF | GCB1 | 0.509 | 0.038 | 13.366 | 0 | 0.693 | 0.435 | 0.584 |
| CF | GCB2 | 0.546 | 0.039 | 14.129 | 0 | 0.743 | 0.471 | 0.622 |
| CF | GCB3 | 0.524 | 0.038 | 13.664 | 0 | 0.712 | 0.448 | 0.599 |
| CF | GCB4 | 0.584 | 0.039 | 14.867 | 0 | 0.795 | 0.507 | 0.661 |
| CF | GCB5 | 0.571 | 0.039 | 14.622 | 0 | 0.777 | 0.495 | 0.648 |
| CF | GCB6 | 0.607 | 0.040 | 15.302 | 0 | 0.826 | 0.530 | 0.685 |
| CF | GCB7 | 0.496 | 0.038 | 13.082 | 0 | 0.674 | 0.421 | 0.570 |
| CF | GCB8 | 0.486 | 0.038 | 12.870 | 0 | 0.661 | 0.412 | 0.560 |
| CF | GCB9 | 0.541 | 0.039 | 14.016 | 0 | 0.736 | 0.465 | 0.616 |
| CF | GCB10 | 0.579 | 0.039 | 14.769 | 0 | 0.788 | 0.502 | 0.656 |
| CF | GCB11 | 0.475 | 0.038 | 12.644 | 0 | 0.647 | 0.402 | 0.549 |
| CF | GCB12 | 0.576 | 0.039 | 14.704 | 0 | 0.783 | 0.499 | 0.652 |
| CF | GCB13 | 0.563 | 0.039 | 14.458 | 0 | 0.766 | 0.487 | 0.639 |
| CF | GCB14 | 0.478 | 0.038 | 12.690 | 0 | 0.650 | 0.404 | 0.551 |
| CF | GCB15 | 0.507 | 0.038 | 13.325 | 0 | 0.690 | 0.433 | 0.582 |
| HF | SF | 1.647 | 0.347 | 4.744 | 0 | 0.855 | 0.966 | 2.327 |

| Factor | Item | Estimate | SE | Z | p | Std.Loading | CI.Lower | CI.Upper |
|---|---|---|---|---|---|---|---|---|
| HF | GF | 0.597 | 0.071 | 8.449 | 0 | 0.513 | 0.459 | 0.736 |
| HF | CF | 0.920 | 0.113 | 8.140 | 0 | 0.677 | 0.699 | 1.142 |

Note. HF = Higher Order Factor; SF = Superstition Factor; GF = Gambling Factor; CF = Conspiracy Factor; GRCS = Gambling Related Cognitions Scale; PDI = Peters Delusion Inventory; SBQ = Superstitious Belief Questionnaire; GCB = Generic Conspiracist Beliefs Scale.

**Table 8**

Factor Loadings for Bifactor CFA

| Factor | Item | Estimate | SE | Z | p | Std.Loading | CI.Lower | CI.Upper |
|---|---|---|---|---|---|---|---|---|
| GB | PDI9 | 0.446 | 0.044 | 10.202 | 0.0000 | 0.447 | 0.361 | 0.532 |
| GB | PDI12 | 0.448 | 0.044 | 10.244 | 0.0000 | 0.448 | 0.362 | 0.534 |
| GB | GCB1 | 0.258 | 0.045 | 5.674 | 0.0000 | 0.258 | 0.169 | 0.347 |
| GB | GCB2 | 0.375 | 0.045 | 8.415 | 0.0000 | 0.375 | 0.287 | 0.462 |
| GB | GCB3 | 0.319 | 0.045 | 7.099 | 0.0000 | 0.320 | 0.231 | 0.408 |
| GB | GCB4 | 0.394 | 0.044 | 8.883 | 0.0000 | 0.394 | 0.307 | 0.481 |
| GB | GCB5 | 0.393 | 0.044 | 8.877 | 0.0000 | 0.394 | 0.307 | 0.480 |
| GB | GCB6 | 0.441 | 0.044 | 10.050 | 0.0000 | 0.441 | 0.355 | 0.526 |
| GB | GCB7 | 0.497 | 0.043 | 11.513 | 0.0000 | 0.497 | 0.412 | 0.581 |
| GB | GCB8 | 0.454 | 0.044 | 10.403 | 0.0000 | 0.455 | 0.369 | 0.540 |
| GB | GCB9 | 0.510 | 0.043 | 11.871 | 0.0000 | 0.511 | 0.426 | 0.594 |
| GB | GCB10 | 0.507 | 0.043 | 11.776 | 0.0000 | 0.507 | 0.422 | 0.591 |

| Factor | Item | Estimate | SE | Z | p | Std.Loading | CI.Lower | CI.Upper |
|---|---|---|---|---|---|---|---|---|
| GB | GCB11 | 0.491 | 0.043 | 11.351 | 0.0000 | 0.491 | 0.406 | 0.575 |
| GB | GCB12 | 0.476 | 0.043 | 10.954 | 0.0000 | 0.476 | 0.391 | 0.561 |
| GB | GCB13 | 0.452 | 0.044 | 10.351 | 0.0000 | 0.453 | 0.367 | 0.538 |
| GB | GCB14 | 0.314 | 0.045 | 6.964 | 0.0000 | 0.314 | 0.225 | 0.402 |
| GB | GCB15 | 0.326 | 0.045 | 7.251 | 0.0000 | 0.326 | 0.238 | 0.414 |
| GB | SBQ1 | 0.771 | 0.038 | 20.065 | 0.0000 | 0.771 | 0.695 | 0.846 |
| GB | SBQ2 | 0.774 | 0.038 | 20.190 | 0.0000 | 0.775 | 0.699 | 0.849 |
| GB | SBQ3 | 0.808 | 0.038 | 21.544 | 0.0000 | 0.809 | 0.735 | 0.882 |
| GB | SBQ4 | 0.635 | 0.041 | 15.427 | 0.0000 | 0.635 | 0.554 | 0.715 |
| GB | SBQ5 | 0.721 | 0.039 | 18.266 | 0.0000 | 0.722 | 0.644 | 0.799 |
| GB | SBQ6 | 0.532 | 0.043 | 12.468 | 0.0000 | 0.532 | 0.448 | 0.616 |
| GB | SBQ7 | 0.756 | 0.039 | 19.509 | 0.0000 | 0.756 | 0.680 | 0.832 |
| GB | SBQ8 | 0.522 | 0.043 | 12.162 | 0.0000 | 0.522 | 0.438 | 0.606 |
| GB | SBQ9 | 0.615 | 0.041 | 14.822 | 0.0000 | 0.616 | 0.534 | 0.696 |
| GB | SBQ11 | 0.752 | 0.039 | 19.364 | 0.0000 | 0.752 | 0.676 | 0.828 |
| GB | SBQ12 | 0.428 | 0.044 | 9.732 | 0.0000 | 0.428 | 0.342 | 0.514 |
| GB | SBQ13 | 0.616 | 0.041 | 14.883 | 0.0000 | 0.617 | 0.535 | 0.697 |
| GB | SBQ14 | 0.736 | 0.039 | 18.808 | 0.0000 | 0.737 | 0.659 | 0.813 |
| GB | SBQ15 | 0.635 | 0.041 | 15.438 | 0.0000 | 0.636 | 0.554 | 0.716 |

| Factor | Item | Estimate | SE | Z | p | Std.Loading | CI.Lower | CI.Upper |
|---|---|---|---|---|---|---|---|---|
| GB | SBQ16 | 0.604 | 0.042 | 14.506 | 0.0000 | 0.605 | 0.523 | 0.686 |
| GB | SBQ17 | 0.502 | 0.043 | 11.635 | 0.0000 | 0.503 | 0.418 | 0.587 |
| GB | SBQ18 | 0.530 | 0.043 | 12.388 | 0.0000 | 0.531 | 0.446 | 0.614 |
| GB | SBQ19 | 0.774 | 0.038 | 20.180 | 0.0000 | 0.774 | 0.698 | 0.849 |
| GB | SBQ20 | 0.798 | 0.038 | 21.114 | 0.0000 | 0.799 | 0.724 | 0.872 |
| GB | SBQ21 | 0.693 | 0.040 | 17.334 | 0.0000 | 0.694 | 0.615 | 0.772 |
| GB | SBQ22 | 0.643 | 0.041 | 15.685 | 0.0000 | 0.644 | 0.563 | 0.723 |
| GB | SBQ23 | 0.642 | 0.041 | 15.643 | 0.0000 | 0.642 | 0.561 | 0.722 |
| GB | SBQ24 | 0.615 | 0.041 | 14.836 | 0.0000 | 0.616 | 0.534 | 0.697 |
| GB | SBQ25 | 0.669 | 0.040 | 16.543 | 0.0000 | 0.670 | 0.590 | 0.749 |
| GB | GRCS3 | 0.482 | 0.043 | 11.131 | 0.0000 | 0.483 | 0.397 | 0.567 |
| GB | GRCS4 | 0.213 | 0.038 | 5.649 | 0.0000 | 0.220 | 0.139 | 0.287 |
| GB | GRCS8 | 0.343 | 0.038 | 9.065 | 0.0000 | 0.358 | 0.269 | 0.417 |
| GB | GRCS9 | 0.147 | 0.039 | 3.777 | 0.0002 | 0.150 | 0.071 | 0.223 |
| GB | GRCS10 | 0.101 | 0.035 | 2.863 | 0.0042 | 0.103 | 0.032 | 0.171 |
| GB | GRCS12 | 0.059 | 0.037 | 1.596 | 0.1105 | 0.059 | -0.013 | 0.131 |
| GB | GRCS13 | 0.269 | 0.039 | 6.878 | 0.0000 | 0.278 | 0.192 | 0.345 |
| GB | GRCS14 | 0.217 | 0.036 | 5.957 | 0.0000 | 0.225 | 0.146 | 0.289 |
| GB | GRCS15 | 0.069 | 0.035 | 2.001 | 0.0454 | 0.070 | 0.001 | 0.137 |

| Factor | Item | Estimate | SE | Z | p | Std.Loading | CI.Lower | CI.Upper |
|---|---|---|---|---|---|---|---|---|
| GB | GRCS18 | 0.295 | 0.038 | 7.840 | 0.0000 | 0.307 | 0.221 | 0.368 |
| GB | GRCS21 | 0.041 | 0.037 | 1.082 | 0.2791 | 0.041 | -0.033 | 0.114 |
| GB | GRCS22 | 0.128 | 0.038 | 3.367 | 0.0008 | 0.131 | 0.054 | 0.203 |
| GB | GRCS23 | 0.347 | 0.040 | 8.689 | 0.0000 | 0.360 | 0.269 | 0.425 |
| SP:S | SBQ4 | 0.304 | 0.042 | 7.278 | 0.0000 | 0.304 | 0.222 | 0.385 |
| SP:S | SBQ5 | 0.342 | 0.037 | 9.301 | 0.0000 | 0.343 | 0.270 | 0.414 |
| SP:S | SBQ8 | 0.284 | 0.046 | 6.138 | 0.0000 | 0.285 | 0.194 | 0.375 |
| SP:S | SBQ9 | 0.426 | 0.041 | 10.360 | 0.0000 | 0.426 | 0.345 | 0.506 |
| SP:S | SBQ15 | 0.318 | 0.042 | 7.660 | 0.0000 | 0.318 | 0.237 | 0.400 |
| SP:S | SBQ16 | 0.310 | 0.043 | 7.216 | 0.0000 | 0.310 | 0.226 | 0.394 |
| SP:S | SBQ17 | 0.406 | 0.046 | 8.889 | 0.0000 | 0.407 | 0.317 | 0.496 |
| SP:S | SBQ18 | 0.431 | 0.044 | 9.707 | 0.0000 | 0.432 | 0.344 | 0.518 |
| SP:S | SBQ22 | 0.235 | 0.042 | 5.589 | 0.0000 | 0.235 | 0.153 | 0.318 |
| SP:S | SBQ23 | 0.280 | 0.042 | 6.733 | 0.0000 | 0.281 | 0.199 | 0.362 |
| SP:S | SBQ24 | 0.447 | 0.041 | 10.950 | 0.0000 | 0.447 | 0.367 | 0.527 |
| SP:G | GRCS1 | 0.608 | 0.042 | 14.466 | 0.0000 | 0.609 | 0.526 | 0.691 |
| SP:G | GRCS2 | 0.537 | 0.043 | 12.447 | 0.0000 | 0.538 | 0.453 | 0.622 |
| SP:G | GRCS4 | 0.540 | 0.040 | 13.383 | 0.0000 | 0.558 | 0.461 | 0.620 |
| SP:G | GRCS5 | 0.684 | 0.041 | 16.816 | 0.0000 | 0.685 | 0.604 | 0.764 |

| Factor | Item | Estimate | SE | Z | p | Std.Loading | CI.Lower | CI.Upper |
|---|---|---|---|---|---|---|---|---|
| SP:G | GRCS6 | 0.601 | 0.042 | 14.256 | 0.0000 | 0.602 | 0.518 | 0.684 |
| SP:G | GRCS7 | 0.607 | 0.042 | 14.424 | 0.0000 | 0.607 | 0.524 | 0.689 |
| SP:G | GRCS8 | 0.465 | 0.039 | 11.935 | 0.0000 | 0.486 | 0.388 | 0.541 |
| SP:G | GRCS9 | 0.532 | 0.042 | 12.777 | 0.0000 | 0.544 | 0.450 | 0.614 |
| SP:G | GRCS10 | 0.660 | 0.040 | 16.531 | 0.0000 | 0.672 | 0.582 | 0.738 |
| SP:G | GRCS11 | 0.611 | 0.042 | 14.545 | 0.0000 | 0.612 | 0.529 | 0.693 |
| SP:G | GRCS12 | 0.632 | 0.041 | 15.392 | 0.0000 | 0.639 | 0.551 | 0.712 |
| SP:G | GRCS13 | 0.462 | 0.041 | 11.355 | 0.0000 | 0.478 | 0.382 | 0.542 |
| SP:G | GRCS14 | 0.578 | 0.040 | 14.608 | 0.0000 | 0.599 | 0.500 | 0.656 |
| SP:G | GRCS15 | 0.695 | 0.040 | 17.536 | 0.0000 | 0.705 | 0.617 | 0.773 |
| SP:G | GRCS16 | 0.528 | 0.043 | 12.205 | 0.0000 | 0.529 | 0.443 | 0.613 |
| SP:G | GRCS17 | 0.539 | 0.043 | 12.505 | 0.0000 | 0.540 | 0.455 | 0.624 |
| SP:G | GRCS18 | 0.502 | 0.039 | 12.738 | 0.0000 | 0.523 | 0.425 | 0.579 |
| SP:G | GRCS19 | 0.604 | 0.042 | 14.332 | 0.0000 | 0.604 | 0.521 | 0.686 |
| SP:G | GRCS20 | 0.693 | 0.040 | 17.114 | 0.0000 | 0.694 | 0.614 | 0.772 |
| SP:G | GRCS21 | 0.618 | 0.041 | 14.894 | 0.0000 | 0.623 | 0.537 | 0.699 |
| SP:G | GRCS22 | 0.567 | 0.041 | 13.720 | 0.0000 | 0.578 | 0.486 | 0.648 |
| SP:G | GRCS23 | 0.378 | 0.040 | 9.365 | 0.0000 | 0.392 | 0.299 | 0.457 |
| SP:C | GCB1 | 0.676 | 0.039 | 17.189 | 0.0000 | 0.677 | 0.599 | 0.753 |

| Factor | Item | Estimate | SE | Z | p | Std.Loading | CI.Lower | CI.Upper |
|---|---|---|---|---|---|---|---|---|
| SP:C | GCB2 | 0.648 | 0.038 | 17.113 | 0.0000 | 0.649 | 0.574 | 0.722 |
| SP:C | GCB3 | 0.656 | 0.039 | 16.923 | 0.0000 | 0.657 | 0.580 | 0.732 |
| SP:C | GCB4 | 0.695 | 0.036 | 19.102 | 0.0000 | 0.696 | 0.624 | 0.767 |
| SP:C | GCB5 | 0.675 | 0.037 | 18.325 | 0.0000 | 0.676 | 0.603 | 0.748 |
| SP:C | GCB6 | 0.701 | 0.035 | 19.944 | 0.0000 | 0.701 | 0.632 | 0.769 |
| SP:C | GCB7 | 0.464 | 0.038 | 12.173 | 0.0000 | 0.464 | 0.389 | 0.538 |
| SP:C | GCB8 | 0.478 | 0.039 | 12.243 | 0.0000 | 0.478 | 0.401 | 0.554 |
| SP:C | GCB9 | 0.531 | 0.037 | 14.503 | 0.0000 | 0.532 | 0.459 | 0.603 |
| SP:C | GCB10 | 0.600 | 0.035 | 16.945 | 0.0000 | 0.601 | 0.531 | 0.670 |
| SP:C | GCB11 | 0.440 | 0.039 | 11.391 | 0.0000 | 0.440 | 0.364 | 0.515 |
| SP:C | GCB12 | 0.617 | 0.036 | 17.142 | 0.0000 | 0.618 | 0.547 | 0.688 |
| SP:C | GCB13 | 0.617 | 0.037 | 16.832 | 0.0000 | 0.617 | 0.545 | 0.689 |
| SP:C | GCB14 | 0.580 | 0.040 | 14.394 | 0.0000 | 0.580 | 0.501 | 0.659 |
| SP:C | GCB15 | 0.622 | 0.039 | 15.810 | 0.0000 | 0.622 | 0.545 | 0.699 |

Note. GB = General Beliefs; SP:S = Superstition Specific Factor; SP:G = Gambling Specific Factor; SP:C = Conspiracy Specific Factor; GRCS = Gambling Related Cognitions Scale; PDI = Peters Delusion Inventory; SBQ = Superstitious Belief Questionnaire; GCB = Generic Conspiracist Beliefs Scale.
